# Asymptomatic SARS-CoV-2 infections tend to occur less frequently in developed nations

**DOI:** 10.1101/2023.12.14.23299954

**Authors:** Shreya Chowdhury, Akshay Tiwari, Ananthu James, Budhaditya Chatterjee, Narendra M. Dixit

## Abstract

Unlike severe infections, asymptomatic infections occur independently of healthcare access and reflect the natural immunity to SARS-CoV-2. What determines their prevalence, *ψ*, and its variation across nations is unknown. We conducted a systematic review of serosurveys performed on samples representative of national populations before vaccination and the emergence of variants. The studies that met our selection criteria together sampled 4,58,489 individuals and yielded estimates of *ψ* in 33 nations. Using random-effects modeling, we found the pooled global *ψ* to be 45.3% (95% CI: 33.6%-57.5%). *ψ* varied widely across nations (range: 6%-96%; *I*^2^=99.7%), highlighting the enormous underlying variation in the natural immunity to SARS-CoV-2. Performing meta-regression with national-level metrics, we found that the human development index (HDI) was negatively correlated with *ψ* (p=10^−13^; *R*^2^=65.5%). More developed nations thus experienced less frequent asymptomatic infections on average. These findings have implications for unraveling the origins of asymptomatic infections and for future pandemic preparedness.

## INTRODUCTION

Asymptomatic infections have been an enigmatic feature of the COVID-19 pandemic^1^. They stand at the favorable end of the spectrum of outcomes following SARS-CoV-2 infection, with the other end being severe respiratory distress and mortality^2^. Their role in driving the spread of the infection was recognized early in the pandemic^3,4^. Significant efforts have since been made to estimate their prevalence^3,5–7^. Yet, unlike severe infections and their associated healthcare burden which were found to vary substantially across nations^8,9^, how the prevalence of asymptomatic infections varies across nations remains unknown. Understanding this variation is important. It has implications for forecasting the course of the pandemic and evaluating intervention strategies^10,11^. It also offers insights into the variation in the natural immunity (as opposed to vaccine-induced immunity) to SARS-CoV-2, a globally new pathogen, across populations: The prevalence of asymptomatic infections, unlike other metrics of disease burden such as the infection fatality ratio^8^, is expected to be robust to differences in health infrastructure and non-pharmaceutical interventions. It is therefore a more reliable indicator of the natural ability of populations to fight the infection, differences in which may arise, for instance, from genetic polymorphisms or differential immunological memory from exposure to related pathogens^12,13^.

Understanding the variation in the prevalence of asymptomatic infections has been challenging for two key reasons. First, epidemiological studies on asymptomatic infections have often resorted to sampling from convenient sources, such as healthcare workers, which introduces biases in the estimates of prevalence^14,15^. For instance, of the 568 positive cases in the New York city jail system, only 2.6% were asymptomatic^16^, whereas of the 61 individuals who tested positive in two large academic health systems in Wisconsin, 88.5% remained asymptomatic^17^. The overall health of individuals in jails is expected to differ vastly from those in academic health systems, potentially resulting in the extreme variation in the prevalence observed across the two samples. Neither of the samples, however, would be representative of the general populations of the two states or of the USA.

Second, factors that could explain the observed variation have not been forthcoming. Meta-analyses have identified factors associated with asymptomatic infections, like the absence of comorbidities, but have not explained the large variation observed^3,5–7^. A recent study discovered a significantly higher frequency of the allele *HLA-B*15:01* in asymptotically infected individuals than in those displaying symptoms, unravelling potential genetic underpinnings of asymptomatic infections^13^. The allele, however, was present in only ∼11% of the asymptomatically infected individuals (and in ∼5% of the symptomatic cases)^13^, leaving a vast majority of the asymptomatic cases unexplained.

Here, we conducted a systematic review of randomized serosurveys, following PRISMA guidelines^18^, to identify studies that reported on asymptomatic infections in samples representative of the general populations of nations. We restricted our review to the early phase of the pandemic when infections were predominantly due to the ancestral SARS-CoV-2 strain and vaccination programs were yet to be initiated. From the studies identified, which spanned nations across continents and socio-economic developmental status, we estimated the extent of variation in the prevalence of asymptomatic infections across nations. We then performed meta-regression to identify key predictors of the prevalence.

## RESULTS

### Systematic review and the global prevalence of asymptomatic infections

We conducted a systematic literature search and identified 12,549 records from four databases that matched our search criteria (4311 from SeroTracker, 4448 from Scopus, 2904 from MedRXiv, and 886 from PubMed, Fig. 1). Applying our inclusion criteria to these records (Fig. 1, Text S1, Methods), we identified 40 articles that offered estimates of the <u>p</u>ercentage of a<u>s</u>ymptomatic infections (*ψ*) from serosurveys on samples representative of the general populations of nations. For identifying asymptomatic infections, serosurveys have an advantage over studies using nucleic acid tests because antibodies last many months after the infection is cleared, allowing a larger time window for detection of infected individuals. They are also, therefore, less likely to encounter a bias from presymptomatic individuals^14^. Among the 40 studies, we found 7 instances where more than one study was from the same nation (Table S1). Interestingly, in all but one of these instances, the different studies from the same nation reported consistent estimates of *ψ* (Table S1), giving us confidence in the estimates. For instance, the two studies from Austria^19,20^ reported *ψ* of 18.0% and 19.7%. This similarity was despite the vastly different seroprevalence–45% and 8.2%, respectively–in the studies, reiterating the notion that *ψ* is an intrinsic property of the populations and is robust to variations in seroprevalence, the latter typically dependent on the state of the pandemic and on non-pharmacological interventions^7^. Given this consistency, we chose one study for each nation–the one with the largest sample size, for its better representativeness of the national population–for further analysis.

**Figure 1:**
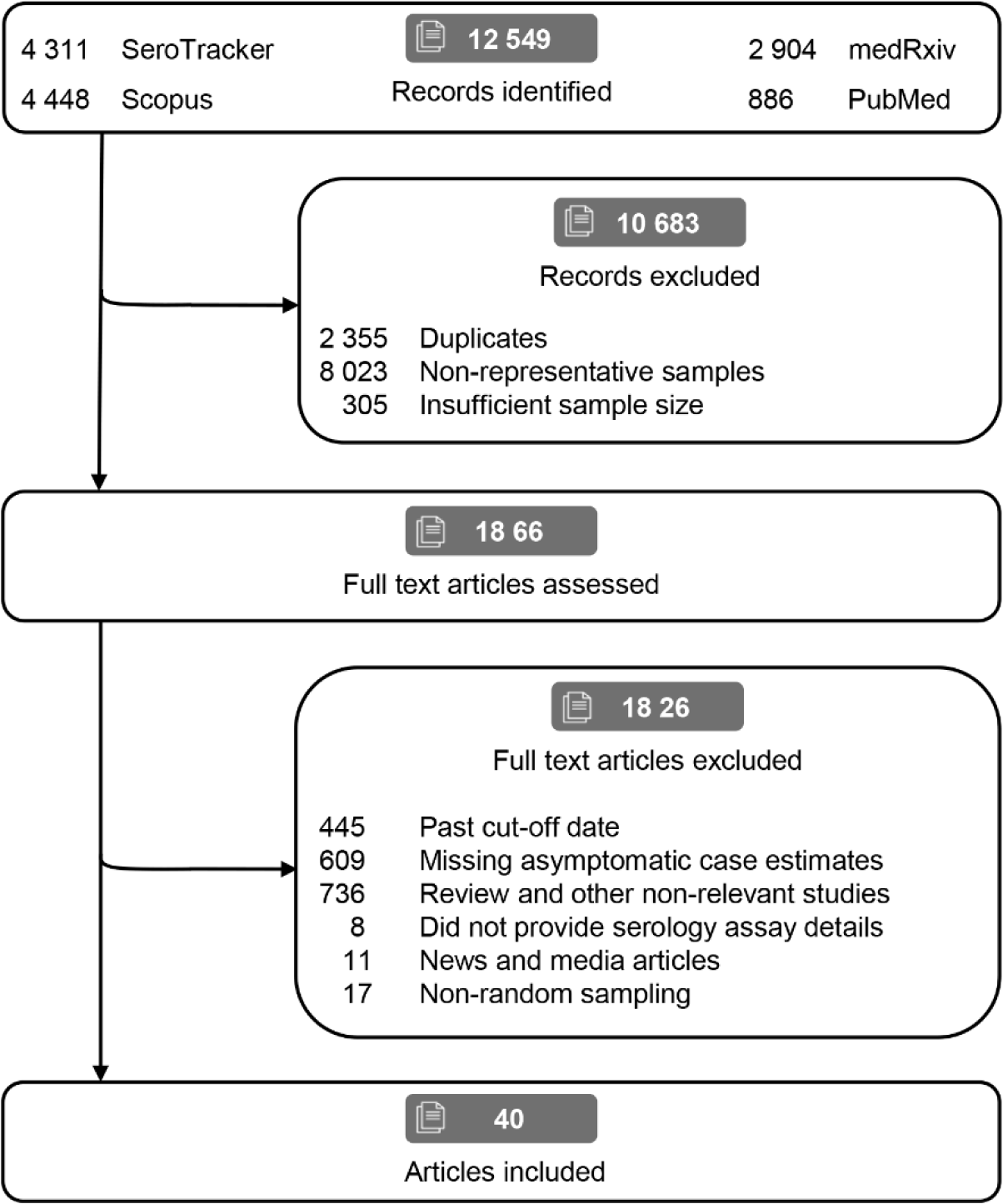
PRISMA flow diagram. The search results and the selection steps leading to the studies included in our meta-analysis.

Thus, we had estimates of *ψ* from 33 studies, one for each of the 33 nations (Table 1; Text S2 provides a brief description of each study). The nations were spread across continents – Asia, the Americas, Europe, and Africa – and across socio-economic developmental status^21^. Of the studies, 21 (63.6%) were nation-wide serosurveys, whereas 6 (18.2%) were conducted at subnational levels and 6 (18.2%) were local serosurveys but using samples representative of the general populations of the respective nations (Table 1). Using a grading scale we developed by adapting the Joanna Briggs Institute (JBI) checklist^22^ (Methods; Text S3), we assessed the 33 studies and found 14 (42.4%) to be of high quality, 14 (42.4%) of moderate quality, and 5 (15.2%) of low quality (Table 1, Table S2). The studies together examined a total of 4,58,489 individuals, of whom 30,086 individuals were seropositive. Of the latter, 15,615 individuals were asymptomatic. To obtain a global average, we estimated the pooled *ψ* across the studies by random effects modeling (Fig. 2, Table S3, Methods). The pooled *ψ* was 45.3% (95% CI: 33.6%-57.5%).

**Figure 2:**
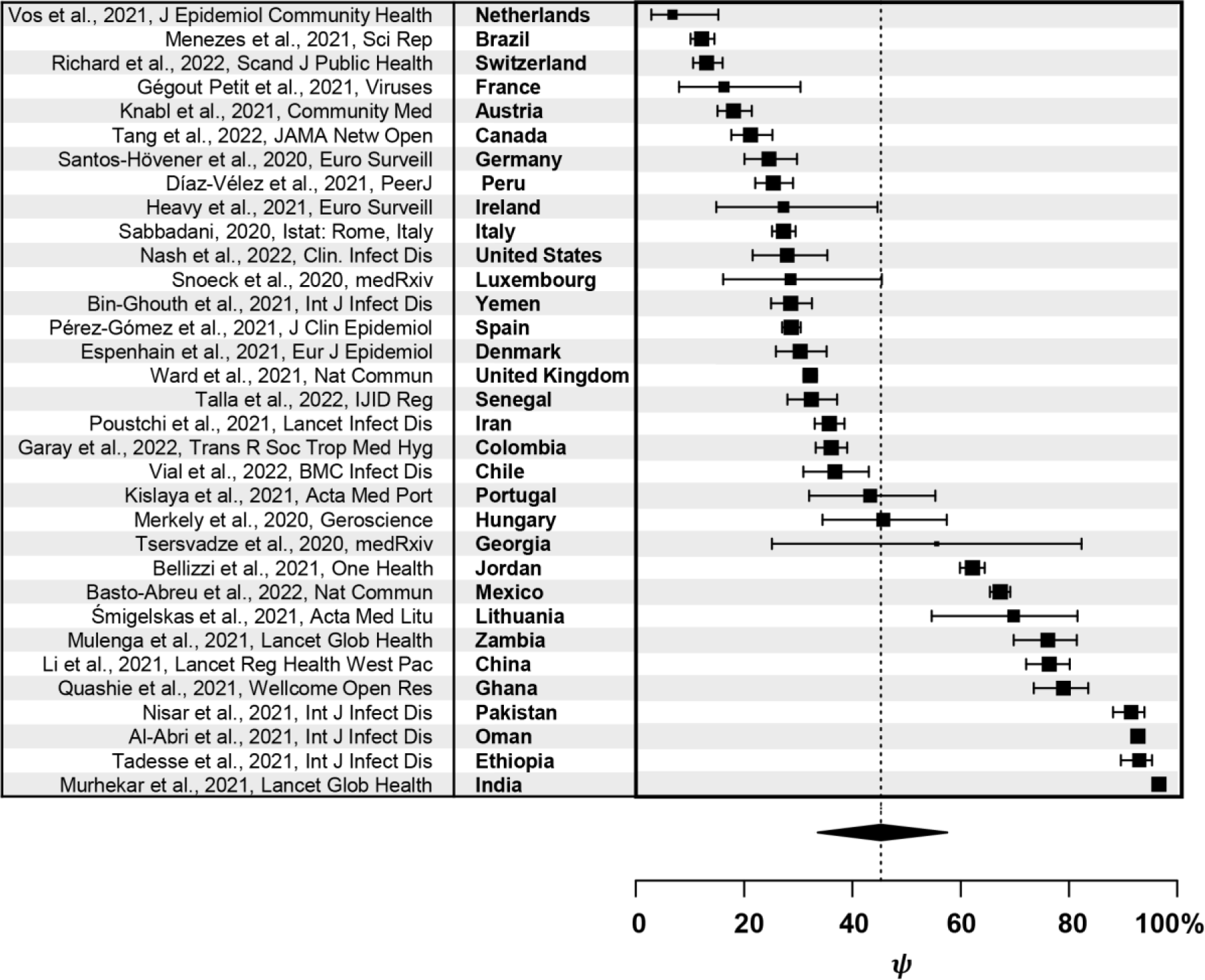
Estimates of *ψ* and its variation across nations. Black rectangles represent *ψ* for the 33 nations studied (see Table 1). Error bars are 95% confidence intervals. The sizes of the rectangles are proportional to the weights assigned to the studies for the random-effects modeling (Methods, Table S3). The vertical dashed line is the pooled estimate of *ψ* (45.3%) with its 95% confidence interval (33.6%-57.5%) shown by the diamond below.

**Table 1:** Summary of the studies included in systematic review. *ψ* is percentage of asymptomatic infections; Q is study quality, with high (H), medium (M), and low (L) categories; G is the geographic scope of the study, with national (N), sub-national (S), and local (L) categories. In cases where the study months but not exact dates were mentioned, the dates have been set to the 15^th^ of the respective months.

| Nation | Study period | Cohort size | Seropositive | Asymptomatic | $\psi$ (95% CI) | Q | G |
| --- | --- | --- | --- | --- | --- | --- | --- |
| Austria<sup>19</sup> | 21-Apr-20 to 27-Apr-20 | 1259 | 566 | 102 | 18.0 (15.1- 21.4) | M | L |
| Brazil<sup>23</sup> | 21-Jun-20 to 24-Jun-20 | 31869 | 849 | 103 | 12.1 (10.1-14.5) | H | N |
| Canada<sup>24</sup> | 01-Dec-20 to 15-Jan-21 | 6955 | 444 | 94 | 21.2 (17.7-25.2) | H | N |
| Chile<sup>25</sup> | 25-Sept-20 to 25-Nov-20 | 2493 | 242 | 89 | 36.7 (31.0-43.0) | H | S |
| China<sup>26</sup> | 10-Apr-20 to 18-Apr-20 | 34857 | 427 | 326 | 76.3 (72.0-80.0) | H | N |
| Colombia<sup>27</sup> | 15-Jul-20 to 15-Nov-20 | 2564 | 1045 | 377 | 36.1 (33.2-39.1) | H | S |
| Denmark<sup>28</sup> | 15-Aug-20 to 11-Dec-20 | 13095 | 369 | 112 | 30.4 (25.9-35.3) | H | N |
| Ethiopia<sup>29</sup> | 24-Jun-20 to 08-Jul-20 | 16932 | 314 | 292 | 93.3 (90.0-95.6) | M | N |
| France<sup>30</sup> | 26-Jun-20 to 24-Jul-20 | 2006 | 43 | 7 | 16.3 (8.1-30.0) | M | L |
| Georgia<sup>31</sup> | 18-May-20 to 27-May-20 | 1068 | 9 | 5 | 55.6 (26.7-81.2) | L | L |
| Germany<sup>32</sup> | 20-May-20 to 09-Jun-20 | 2203 | 249 | 61 | 24.5 (19.6-30.2) | M | L |
| Ghana<sup>33</sup> | 27-Jul-20 to 14-Sep-20 | 1305 | 252 | 199 | 79.1 (73.7-83.7) | L | N |
| Hungary<sup>34</sup> | 01-May-20 to 16-May-20 | 10474 | 70 | 32 | 47.1 (35.9-58.6) | M | N |
| India<sup>35</sup> | 18-Aug-20 to 20-Sep-20 | 29082 | 3135 | 3029 | 96.6 (95.9-97.2) | M | N |
| Iran<sup>36</sup> | 17-Apr-20 to 02-Jun-20 | 3530 | 1164 | 416 | 35.7 (33.0-38.5) | H | N |
| Ireland<sup>37</sup> | 22-Jun-20 to 16-Jul-20 | 1733 | 33 | 9 | 27.3 (15.1-44.2) | M | S |
| Italy<sup>38</sup> | 25-May-20 to 15-Jul-20 | 64660 | 1617 | 441 | 27.3 (25.2-29.5) | H | N |
| Jordan<sup>39</sup> | 27-Dec-21 to 06-Jan-21 | 5044 | 1723 | 1071 | 62.2 (59.9-64.5) | L | N |
| Lithuania<sup>40</sup> | 10-Aug-20 to 10-Sep-20 | 3087 | 43 | 30 | 69.0 (54.1-80.8) | H | N |
| Luxembourg<sup>41</sup> | 15-Apr-20 to 05-May-20 | 1807 | 35 | 10 | 28.6 (16.3-45.0) | H | N |
| Mexico<sup>42</sup> | 15-Aug-20 to 15-Nov-20 | 9640 | 2400 | 1615 | 67.3 (65.4-69.1) | H | N |
| Netherlands<sup>43</sup> | 31-Mar-20 to 11-May-20 | 3147 | 74 | 5 | 6.8 (2.9-14.9) | M | N |
| Oman<sup>44</sup> | 12-Jul-20 to 08-Dec-20 | 17457 | 3841 | 3562 | 92.7 (91.8-93.4) | M | N |
| Pakistan<sup>45</sup> | 15-Apr-20 to 22-Aug-20 | 3005 | 364 | 333 | 91.5 (88.2-94.0) | M | L |
| Peru<sup>46</sup> | 24-Jun-20 to 10-Jul-20 | 2010 | 595 | 151 | 25.4 (0.22-0.29) | M | L |
| Portugal<sup>47</sup> | 21-May-20 to 08-Jul-20 | 2301 | 67 | 29 | 44.0 (32.8-55.9) | H | N |
| Senegal<sup>48</sup> | 25-Oct-20 to 26-Nov-20 | 1422 | 398 | 129 | 32.4 (28.0-37.1) | M | N |
| Spain<sup>49</sup> | 27-Apr-20 to 22-Jun-20 | 61092 | 2669 | 766 | 28.7 (26.1-31.4) | M | N |
| Switzerland<sup>50</sup> | 06-Apr-20 to 30-Jun-20 | 8344 | 590 | 77 | 13.1 (10.6-16.1) | H | S |
| UK<sup>51</sup> | 20-Jun-20 to 13-Jul-20 | 105651 | 5544 | 1785 | 32.2 (31.0-33.4) | H | N |
| USA<sup>52</sup> | 15-May-20 to 15-Jan-21 | 4510 | 161 | 45 | 28.0 (21.6-35.4) | L | N |
| Yemen<sup>53</sup> | 15-Nov-20 to 15-Dec-21 | 2001 | 549 | 157 | 28.6 (0.25-0.33) | L | S |
| Zambia<sup>54</sup> | 04-Jul-20 to 27-Jul-20 | 1886 | 205 | 156 | 76.2 (69.9-81.5) | M | S |

### Variation of the prevalence of asymptomatic infections across nations

Interestingly, the estimates of *ψ* revealed an enormous variation across nations (Table 1, Fig. 2). The Netherlands had the lowest *ψ*, 6%, and India the highest, 96%. 7 nations had *ψ* between 75% and 100%, 4 between 50% and 75%, 16 between 25% and 50%, and 6 below 25%. We estimated confidence intervals on *ψ* using the Wilson score interval (Methods). The intervals closely matched estimates where available (Table S4). Small sample sizes often led to large confidence intervals, as with Georgia (Fig. 2). Nonetheless, the data yielded an inconsistency index (*I*^2^) of 99.7% (Methods), implying extremely high inter-nation heterogeneity, not attributable to sampling variations.

This wide variation in *ψ* was intriguing. It suggested that substantial differences exist in the natural immunity of the populations in different nations to the infection. To elucidate plausible origins of this variation, we next performed meta-regression.

### Meta-regression and the human development index

Key clinical and demographic correlates implicated in severe COVID-19 infections include age, comorbidities, and prior exposure to related pathogens^55^. While similar factors are implicated in asymptomatic infections^12^, population level predictors of *ψ* are yet to be identified. Here, we therefore assessed whether the above predictors of severe infections could also predict the variation in *ψ*. A population level metric of the prevalence of comorbidities is lacking. The largest cause of mortality in adults across the globe is cardiovascular disease (CVD)^56^. We therefore used the prevalence, or rate, of CVD per 10^5^ individuals (CVDR) as a proxy for the prevalence of comorbidities. While severe infections tend to increase with age, asymptomatic infections within a nation seemed to be much less sensitive to age, barring children and the elderly^23,24,49,51^ (Fig. S1). Besides, intriguingly, some studies have observed a higher prevalence of asymptomatic infections in the elderly^24,51^. We therefore considered the demographic median age (DMA) of each nation as a metric representative of the ages of the respective national populations. Prior exposure to related pathogens is difficult to quantify at the population level. We reasoned, following recent evidence, that the overall prevalence of infections may be a good indicator of the exposure to related pathogens, including circulating coronaviruses, and hence of the pre-existing immunity to SARS-CoV-2^12^. The overall prevalence of infections is often dependent on the development status of a nation, as observed, for instance, with lower respiratory tract infections (LRIs) over the last several decades^57^. LRIs were more prevalent in less developed nations^57^. The human development index (HDI) is a widely used marker of the socio-economic developmental status of nations^21^. The higher the HDI, the more developed is a nation. We therefore considered HDI as a proxy for prior exposure to related pathogens. We collected national-level estimates of these predictors for the nations included in our analysis (Table S5).

Before performing meta-regression, we assessed whether the predictors, CVDR, DMA, and HDI, were correlated. We found that the predictors were all highly correlated (Fig. S2). For regression, we therefore constructed models with the predictors chosen one at a time (Methods). To identify influential observations, including which in the regression would substantially adversely affect the model performance, we performed a detailed influential case diagnosis for each model (Methods, Fig. S3). This involved performing meta-regression with each model by leaving out one observation at a time and evaluating the model performance. For the model with CVDR as predictor, we found that *ψ* for Lithuania was an influential observation. For the model with DMA, *ψ* for Senegal and Yemen were influential observations. For the model with HDI, the influential observations were *ψ* for Senegal, Yemen, and Oman. For each model, we therefore excluded the respective influential observations and performed meta-regression using logit-transformed *ψ*.

The model with HDI as the predictor emerged as the best model (Table S6). The best-fit showed that HDI was strongly negatively correlated with *ψ* (*p* = 4 × 10^−13^; Fig. 3). Examining nations based on the United Nations guidelines for HDI-based categorization of developmental status^58^, we found that nations with low (<0.55) and medium (0.55– 0.699) HDI had a much higher *ψ* compared to their high (0.7–0.799) and very high (0.8– 1) HDI counterparts (Fig. 3). Remarkably, HDI explained nearly 65.5% of the variation in *ψ* observed. Models with DMA or CVDR as predictors explained much less of the variation, ∼42.4% and ∼24.8%, respectively (Table S6; Fig. S4).

**Figure 3:**
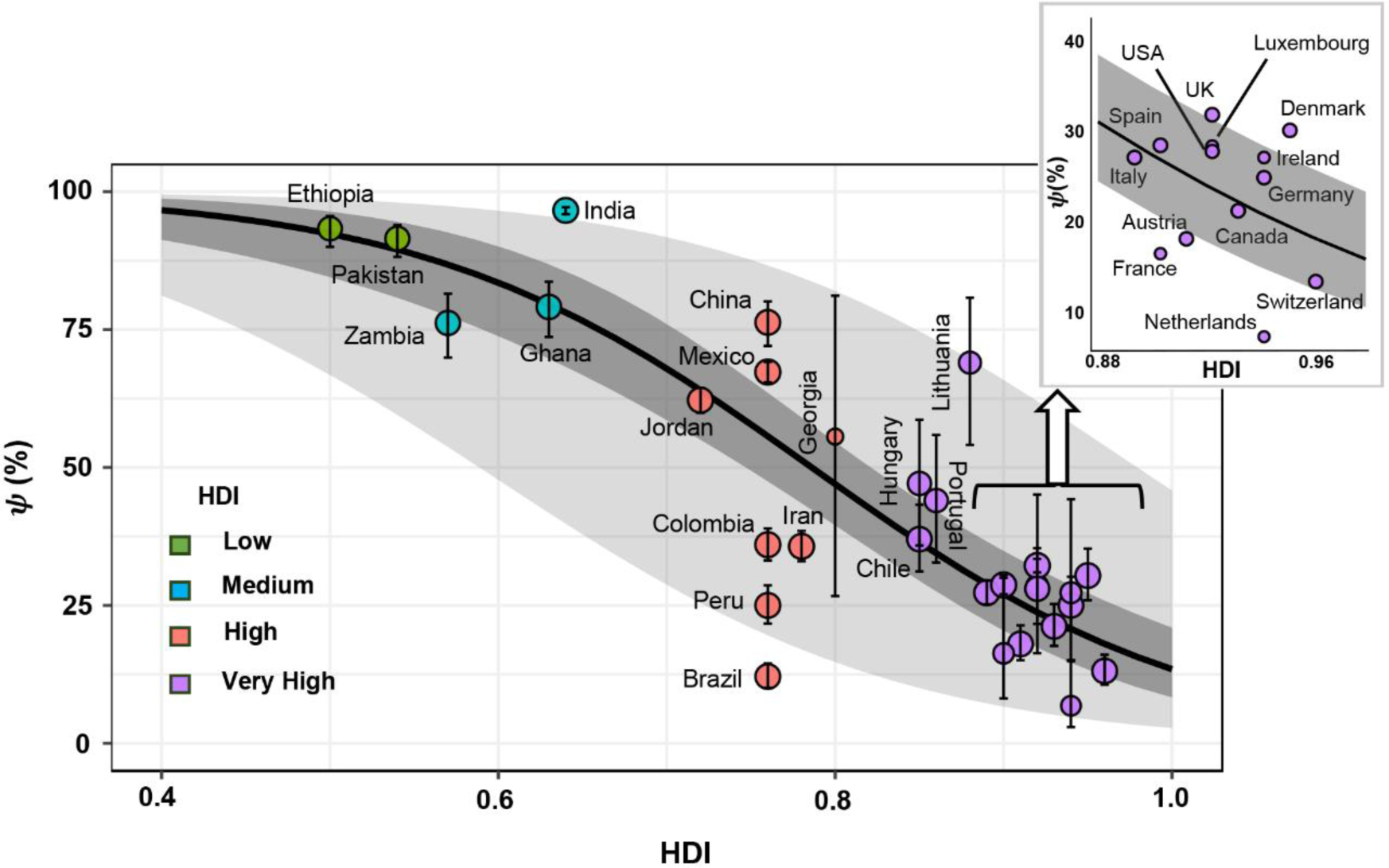
HDI is a robust predictor of *ψ*. Best-fit meta-regression line (solid line) showing the negative correlation between *ψ* and HDI, overlayed on the estimates of *ψ* from 30 nations (symbols) (omitting the 3 influential observations). The dark ribbon is the 95% confidence interval and the light ribbon the 95% prediction interval. The symbol sizes are proportional to the weights of the studies in the meta-regression (see Methods). The symbols are color-coded according to the HDI categories indicated. Error bars are the same as in Fig. 2. Regression coefficients and statistics are in Table S6.

We ruled out the influence of potential confounding factors. First, although the studies used antibody tests with high specificities, they had varying sensitivities (Table S7). We found that *ψ* was uncorrelated with the sensitivity of the tests (Fig. S5), ruling out any confounding effect from the assay variations. The studies also used distinct symptom sets in the questionnaires (Fig. S6). A study using fewer symptoms may over-estimate asymptomatic cases. We found that *ψ* was uncorrelated with the number of symptoms used (Fig. S7). We noticed that most nations used fever and cough as part of their symptom set. It is rare that other symptoms are experienced without also experiencing fever or cough^59^. Consequently, the other symptoms may not significantly increase the chances of detecting symptomatic cases, explaining the lack of a correlation between *ψ* and the number of symptoms. Finally, we assessed whether distinct age distributions in the nations included could confound our inferences. In particular, the presence of children or the elderly could skew estimates of *ψ*. Although several studies included children, children accounted for only a small fraction of the seropositive cases (Table S8). Their presence would thus not affect estimates of *ψ* substantially. The greater prevalence of the elderly in more developed nations could confound the association of *ψ* with HDI. We therefore performed meta-regression with the percentage of the population aged >65 years in each nation as the predictor. The predictor explained only 37% of the variation in *ψ* observed (Fig. S8, Table S9), rendering it a much poorer predictor than HDI (and median age). HDI was thus a robust predictor of *ψ*.

## DISCUSSION

Remarkable differences have been observed in the burden of COVID-19 across nations^8,9^. The differences arise not only from differences in health infrastructure, non-pharmaceutical interventions, and demographics across nations, but also from potential natural variations in immunity. While the former factors have been widely studied^8,9^, natural variations have remained elusive. Here, we recognized that the prevalence of asymptomatic infections, *ψ*, serves as a reliable measure of the natural immunity of a population to SARS-CoV-2 and carefully collated and assessed its variation across nations. Our findings revealed the enormous variation in the natural immunity to SARS-CoV-2 across nations.

An intriguing question that follows is what determines this variation. Studies have attempted to assess whether factors responsible for severe infections^8,9,60^ also explain asymptomatic infections^61^. Genetic factors too have been assessed^13^. However, no studies so far have considered metrics such as *ψ* quantified across nations. The origins of the variation at the population level have therefore remained largely unknown^61^. Here, we found that HDI was the best of the predictors of *ψ* we studied, explaining over 65% of its variation. We contrast this with the genetic polymorphisms identified, which leave over 90% of the asymptomatic cases unexplained^13^.

HDI is a marker of the overall development of a nation, determined as a composite of three measures, namely, life-expectancy at birth, years of schooling, and per capita income^21^. That *ψ* was negatively correlated with HDI implied that more developed nations experienced fewer asymptomatic infections on average and hence, presumably, had weaker natural immunity. Because HDI is a composite metric, a precise, causal relationship between HDI and natural immunity is difficult to establish. In more developed nations, due to long-term lifestyle changes, including the practice of hygiene and the use of antibiotics, the prevalence of infections in general is lower^57,62^. We speculate that this may be responsible for the weaker immunity to SARS-CoV-2, possibly due to altered microbiota^63^ and/or weaker trained immunity^64^. In addition, the associated lower prior exposure to circulating coronaviruses may limit asymptomatic infections^65^.

We do not rule out the role of other factors associated with severe infections, such as age and comorbidities, in establishing asymptomatic infections. Indeed, in our meta-regression, median age (DMA) and the prevalence of cardiovascular disease (CVDR) explained ∼42% and ∼25% of the variation in *ψ*. However, because these predictors were strongly correlated with HDI, their specific contributions, over those attributable to their correlation with HDI, remain to be delineated.

Severe infections increase significantly with age^8,9^. Asymptomatic infections may thus be expected, conversely, to decrease with age. We found, surprisingly, from age-stratified data from four nations that *ψ* within a nation was only weakly dependent on age (Fig. S1). Furthermore, in the UK and Canada, *ψ* increased with age (Fig. S1). An explanation of these counter-intuitive trends is lacking^1^. Nonetheless, while age-stratified data for the other nations we studied was not available, the data above suggests a limited role for age compared to HDI in explaining the variation in *ψ*.

We recognize that deviations in *ψ* from the mean trends predicted by HDI exist. For instance, China and Brazil had identical HDIs but widely different estimates of *ψ* (Fig. 3). Factors currently unknown that could explain the remaining ∼35% of the variation in *ψ* not explained by HDI may help reconcile these deviations.

Our finding that HDI is the predominant factor predicting *ψ* may inform ongoing studies aimed at elucidating the mechanistic origins of asymptomatic infections. We recall that a key outcome of COVID-19 vaccination has been to render potentially symptomatic infections asymptomatic, which most approved vaccines achieved with high efficacies^66,67^. Vaccination studies have offered correlates of protection, such as the antibody titre, that may help predict the likelihood of an infection remaining asymptomatic^66,67^. Interestingly, the genetic polymorphisms identified recently too point to pre-existing immunity, specifically T-cell immunity, possibly triggered by pre-pandemic exposure to circulating coronaviruses, as associated with asymptomatic infections^13^. Future studies may assess how differences in HDI may lead to differences in such correlates of protection across nations, leading to a better understanding of the natural immunity underlying asymptomatic infections and the variation in *ψ*.

Our study also has implications for understanding the spread of the pandemic and for intervention strategies. Asymptomatic infections have been estimated to have been responsible for nearly a quarter of all transmission events^4^. Transmission by asymptomatic individuals is hard to contain because the individuals are not readily identified. Modeling studies have therefore used estimates of *ψ* to forecast the course of the pandemic and evaluate intervensions^10,11^. Our findings may enable such models to make more accurate predictions. The larger *ψ* in less developed nations, for instance, would imply that in these nations, especially in the more populous ones like India, a much larger share of the transmission events may have been due to asymptomatic individuals, exacerbating the difficulty in containing the pandemic. This may have led, compounded by the lack of adequate healthcare access, to the increased mortality in these nations^8^. Quantifying these differences, especially in the context of emerging variants, would be important for future pandemic preparedness.

## ONLINE METHODS

### Systematic Review

#### Search Strategy

We systematically reviewed studies on seroprevalence, focusing on the proportion of asymptomatic SARS-CoV-2 infections (*ψ*). This review adhered to the Preferred Reporting Items for Systematic Reviews and Meta-Analysis (PRISMA) guidelines^18^. We followed a primary search strategy to extract serosurvey studies from the open access SeroTracker^68^ database platform. Our secondary search involved comprehensive screening of COVID-19 and SARS-CoV-2 compendiums from PubMed, SCOPUS and MedRxiv without language restriction. We employed the terms ‘COVID-19,’ ‘SARS-Cov-2,’ ‘seroprevalence,’ ‘serological test,’ and ‘asymptomatic’ as search keywords when querying the databases. We attempted to mitigate possible publication bias by assessing both published articles and unpublished literature such as grey literature, preprints, institutional reports. We omitted news and media reports. All but four source articles from where we extracted the data for our meta-regression were published in peer-reviewed journals (Table 1). Of the studies not peer-reviewed, three were preprints (the articles on Georgia^31^, Ghana^33^, and Luxembourg^41^), and one was a report from governmental sources (Italy^38^). We included studies conducted between March 11, 2020, when WHO declared the COVID-19 pandemic, and December 8, 2020, when the first vaccine was administered outside clinical trials^69^. We additionally included the studies from USA^52^, Canada^24^, and Jordan^39^ because, although they exceeded the above timeframes, either no vaccination had begun in these nations till the end of the study period or the vaccinated individuals were omitted from the reported serosurvey data (see Text S2).

This review was not registered. Two researchers (AT and SC) reviewed the title, abstracts, and full texts of the articles independently. Disagreements were resolved by three other reviewers (AJ, BC, and NMD). The researchers conducted initial searches from November 22, 2022, to December 13, 2022, and continued to perform monthly searches thereafter until September 20, 2023. Data collection did not involve any automated processes.

#### Selection Criteria

Asymptomatic individuals are those who become infected but never develop or perceive any symptoms during the entire course of infection^14^. We chose retrospective cross-sectional serosurveys that determined the proportion of asymptomatic infection through interviews and questionnaires, collecting data on symptoms reported either at the time of blood sample collection or during a prior period. The study window chosen (see above), which lasted under a year of the onset of the epidemic in the respective settings, minimized the chances of seroreversion^14^.

While screening available studies, we identified those that provided accurate estimates of asymptomatic cases and were conducted on participants representing the national populace, by methods such as household or community sampling. We ensured a sample size n > 500. When investigations of such scale were not available for a nation, we included studies that reported the seroprevalence in provinces^26,45^, counties^37^, regions^42^, governates^39^, major cities and towns^20,29,32,36,40^, districts^54^, or municipalities^27,30,35^, with populations that were representative of the general population of the nation.

Subsequently, the complete texts of the screened studies underwent evaluation (Fig. 1, Text S1). Serosurveys conducted on vaccinated individuals or after the emergence of variants were excluded. Studies that lacked reporting of the total number of individuals tested, details on seroprevalence, or the number/percentage of asymptomatic individuals among the seropositive participants were also excluded. Studies that lacked details of serology tests were excluded. Further, we excluded review articles and findings sourced from news and media outlets. Finally, studies that did not ensure random sampling of their participants were excluded. Collated in this manner, we ensured inclusion of the most reliable evidence coming from large-scale cross-sectional seroprevalence studies with representative samples that included data from antibody-based testing and identified population-level asymptomatic proportions in a nation.

#### Data Extraction and Quality Assessment

From each study, data of nation, sample frame (group of interest), study duration including sampling start and end date, geographical scope of the study (national, regional, local, sub-local), age of participants, number of tested individuals, number of confirmed COVID-19 infections, number of asymptomatic infections, comorbid conditions, participation rate, type of serological assay, test kit name and details, sensitivity and specificity of the serological assay, number of symptoms recorded in the questionnaire, source type, first author, and date of publication were extracted. Two reviewers (AT and SC) performed the quality assessment independently. Disagreements were resolved by three other reviewers (AJ, BC, and NMD). Because the outcome of our interest was the proportion of asymptomatic infections and not seroprevalence, we adapted the Joanna Briggs Institute (JBI) checklist^22^ to develop our own quality assessment tool and grading scale (Text S3). Our quality assessment tool assessed the domains of symptom assessment, reliability of the serological assays and questionnaires, and the sampling method to ensure representativeness at the national level (Table S2). The studies were classified as low, moderate, or high quality following the grading scales developed a priori (Text S3). Neither the data extraction nor the quality assessment involved any automated process.

#### Data on Predictors

We collated national-level data on demographic median age, population age distribution, prevalence of cardiovascular disease, and human development index (Table S5). All the data were for 2019.

### Statistical analysis and meta-regression

We used *ψ* as the outcome measure. We calculated confidence intervals on *ψ* using the Wilson score interval^70^. For computing the pooled estimate of *ψ*, we logit transformed the reported estimates, 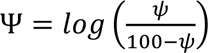, and assigned individual weights to each study using the inverse variance method^71^. We used the Paule and Mandel method^72^ implemented in the Metafor^73^ package in R for parameter estimation. To quantify the heterogeneity, we evaluated the *I*^2^ metric^74^. We calculated the Spearman’s correlation coefficient to evaluate the correlations between the predictors, CVDR, DMA, and HDI. We performed meta-regression using the predictors one at a time. To identify outliers and influential observations, we estimated studentized residuals and performed meta-regression by leaving out one observation at a time. An observation was classified as influential if the absolute values of the studentized residuals for the observation were larger than 3 and/or leaving it out resulted in an increase in *R^2^* greater than 5 percentage points. We compared the resulting models using *R^2^*. We performed additional tests to ascertain the robustness to potential confounding factors (see Results).

## Data Availability

All data produced in the present work are contained in the manuscript.

## ACKNOWLEDGEMENTS

We thank Shreyas Joshi for insightful discussions. BC was supported by the C. V. Raman postdoctoral fellowship of the Indian Institute of Science.

## Notes

### Competing Interest Statement

The authors have declared no competing interest.

