## Supplementary Materials for "Asymptomatic SARS-CoV-2 infections tend to occur less frequently in developed nations"

This file includes:

Text S1: Inclusion and exclusion criteria

Text S2: Description of the studies included in meta-analysis

Text S3: Description of critical appraisal process

Table S1: Estimates of $\psi$ across independent studies from specific nations

Table S2: Critical appraisal of the studies included in the systematic review

Table S3: Weights assigned to studies in the random-effects model

Table S4: Reported versus calculated confidence intervals on $\psi$

Table S5: Data of the predictors

Table S6: Model characteristics and parameter estimates

Table S7: Sensitivity and specificity of the assays used

Table S8: Reported seropositivity in children

Table S9: Model characteristics and parameter estimates with age≥65 y as predictor

Figure S1: Age stratified $\psi$ in studies of Brazil, Canada, Spain, and UK

Figure S2: Correlation between predictors

Figure S3: Influential case diagnostics

Figure S4: Meta-regression with other predictors

Figure S5: Robustness to assay sensitivity

Figure S6: Symptom sets

Figure S7: Robustness to symptom sets

Figure S8: Meta regression of $\psi$ on age $\geq$ 65 y

### **Text S1:** Inclusion and exclusion criteria

We employed the following inclusion criteria:

1. Studies that reported seroprevalence data based on a nationally representative samples
2. Studies that involved random selection of participants
3. Studies that reported asymptomatic infections using a questionnaire on serosurveyed individuals
4. Studies with sample size >500

We subjected the studies selected above to the following exclusion criteria:

1. Studies conducted outside March 11, 2020 to December 8, 2020.
2. Reviews and non-serosurveys
3. Studies that failed to provide details of the serological test assays
4. News and media articles

**Text S2: Description of the studies included in meta-analysis.**

**I. Austria[1] –** The seroprevalence study was performed in Ischgl/Tyrol, a tourism hub in Austria. Conducted from April 21 to 27, 2020, this cross-sectional epidemiological survey encompassed all residents, regardless of their age or gender. Participants' blood samples underwent screening for anti-SARS-CoV-2-S1-protein IgA and IgG using commercially available ELISA tests. Out of 1867 inhabitants in Ischgl, 1473 individuals from 478 households, representing various age groups, were tested. Among them, 566 individuals tested positive. 18% of those who were seropositive reported no symptoms.

**II. Brazil[2] –** In this nationwide seroprevalence study, undertaken during May and June, 2020, the most populated municipalities of all 133 intermediate regions, which cover each of the 26 states as well as the federal district of the nation, were selected for sample collection. Households were selected randomly within these municipal areas and one individual per household was selected randomly for collection of capillary blood sample by finger pricking. Survey results from 21-24 June, 2020 are reported. A total of 31,869 individuals consented for the sample collection procedure and responded to the questionnaire. The samples were tested with a lateral flow immunoassay to detect the presence of anti-SARS-CoV-2 IgG and IgM antibodies. 849 participants were seropositive. Among them 12.1% did not report any symptoms.

**III. Canada[3] –** This study recruited individuals enlisted by a public polling organization from across the regions in Canada. All participants were adults (>18 years). The authors stratified the cohort for age and sex, to match the national demographic distribution. The study was undertaken in two different phases on the same cohort: the first one lasted from May to September 2020 and the second from December 2020 to March 2021. We used the data from the second phase because a higher number of seropositive individuals was reported and they were more homogenously distributed across the nation than from the first phase. The participants were invited via email and requested to respond to a questionnaire. Once they consented to provide blood samples, dried blood sample collection kits were dispatched to them via postal service. The participants self-collected the sample and returned it to the authors. A chemiluminescence assay was used to detect IgG antibodies against the S and N viral proteins. In the second phase, 6955 respondents provided dried blood samples. Among them 444 were seropositive. Among the latter, 94 (21.2%) participants declared not experiencing symptoms. The second phase survey ended in January 2021. The authors removed vaccinated individuals from the second phase data.

**IV. Chile[4] –** Between September and November, 2020, a population-based serosurvey was conducted among residents aged seven years and older in three urban areas situated in the central zone of Chile: Santiago, Talca, and Coquimbo–La Serena. Participants were selected through a two-stage stratified sampling process. Those who consented to participate completed an on-site questionnaire. SARS-CoV-2 antibodies were detected using ELISA. In cases where venipuncture failed, was contraindicated, or for young children who refused venipuncture, a point-of-care rapid test was employed. Of the 2493 participants, 10.4% (95% CI 7.8-13.7%) were seropositive. Among the latter, approximately 36% were asymptomatic.

**V. China[5] –** The serosurvey was conducted in the city of Wuhan and the corresponding province, as well as six other provinces where the highest prevalence of the infection was recorded. The drive for the collection of the samples continued from 10$^{th}$ to 18$^{th}$ April, 2020. The authors randomly chose counties within the study areas, and then randomly chose communities within those counties, and finally randomly selected households within the communities. If the willing participants were not present in the survey area from December to March 2020 for more than two weeks, they were excluded. All participants were above 1 year of age. The consenting participants were interviewed based on a structured questionnaire, seeking personal data and COVID-19 symptomatology. A total of 34,857 participants provided blood samples. The samples underwent three tiers of testing. A lateral flow immunoassay was followed by a chemiluminescence enzyme assay to detect seropositive samples. Positive results were confirmed with a microneutralization assay. 427 individuals were confirmed to be seropositive by the microneutralization assay. Among the seropositive individuals, 76.3% were asymptomatic.

**VI. Colombia[6] –** Eight municipalities with the largest populations in a department (administrative region) of the Colombian Caribbean were selected for the seroprevalence study spanning July to November 2020. A total of 2564 individuals consented and provided blood samples and responded to the questionnaire. An ELISA was used to detect IgG, IgA and IgM antibodies against N protein of SARS-CoV-2. 1045 individuals were seropositive. Of them, 377 (36.1%) were asymptomatic.

**VII. Denmark[7] –** A nationwide serosurvey in Denmark was undertaken between August to December, 2020, in three rounds. Participants were randomly selected from the national civil registry and subsequently invited to donate blood samples. A total of 13,095 individuals provided samples and answered the structured questionnaire. An ELISA was used to detect anti-SARS-CoV-2 IgG antibodies in the serum. 369 individuals were seropositive, among whom 30% declared not to have experienced symptoms.

**VIII. Ethiopia[8] –** A nationwide seroprevalence study was conducted from 24$^{th}$ June to 8$^{th}$ July, 2020 in 14 major urban areas across Ethiopia. Households were selected randomly and venous blood samples from all household members above 15 years of age were collected. The samples were tested with a chemiluminescent microparticle immunoassay to detect IgG antibodies. 16,932 got tested and completed the questionnaire. 313 individuals tested seropositive. Among them, 21 were reported to have had symptoms.

**IX. France[9] –** Individuals were randomly selected from the electoral lists of the city Nancy and invited along with the members of their households to donate blood samples for the serosurvey. Samples from a total of 2006 individuals were collected from 26$^{th}$ June to 24$^{th}$ July 2020. They also responded to a questionnaire. The samples were tested using an ELISA to detect IgA, IgG, and IgM antibodies against SARS-CoV-2. 43 individuals were seropositive, among whom 16.3% were asymptomatic.

**X. Georgia[10] –** Participants were recruited across the national capital region of Georgia using a respondent dependent sampling strategy. The samples were collected from the participants from 18$^{th}$ to 27$^{th}$ May 2020. A total of 1068 individuals provided blood samples and answered questions in a short interview, about personal data and symptomatology. A lateral flow immunoassay was used by the authors to detect SARS-CoV-2 specific IgG and IgM antibodies. 9 were seropositive, and among them 5 (56%) reported no symptoms.

**XI. Germany[11] –** Randomly selected adults from the township of Kupferzell were recruited for the serosurvey. A total of 2203 individuals provided their consent to the collection of blood and answered a questionnaire. The sample collection drive lasted from 20$^{th}$ May to 9$^{th}$ June, 2020. The samples were tested with an ELISA to detect anti-SARS-CoV-2 IgG antibodies. 249 individuals were seropositive. Among them, 24.5% individuals reported not to have experienced any symptoms.

**XII. Ghana[12] –** The first phase of the seroprevalence study from Ghana lasted from 27$^{th}$ July to 14$^{th}$ September, 2020. Blood samples of 1,305 randomly picked individuals aged 4 years and above from selected locations across Ghana were tested with a lateral flow rapid diagnostic test kit, which detects IgG and IgM antibodies against SARS-CoV-2. While sampling, individuals experiencing symptoms, such as fever, were excluded. Among all tested individuals 19% were seropositive. Among the seropositive individuals 20.9% declared symptoms.

**XIII. Hungary[13] –** In a nationwide serosurvey, participants older than 14 years were randomly selected from randomly selected households in 489 settlements (localities). The settlements were selected across the nation from which one or more confirmed COVID-19 cases were reported till then. A total of 10474 participants provided consent for the collection of blood and other samples and responded to a questionnaire. The sample collection drive lasted from 1$^{st}$ to 16$^{th}$ May, 2020. The authors used a chemiluminescent microparticle immunoassay to detect anti-SARS-CoV-2 IgG antibodies. They also tested the nasopharyngeal and oropharyngeal swabs via RT-PCR. 70 individuals tested positive on both. Among them, 37 participants reported symptoms.

**XIV. India[14] –** The nationwide seroprevalence study included households from 700 villages and wards (electoral subdivisions) from 70 districts encompassing 21 states in the nation. The households were randomly selected in the study areas and the household members were invited to participate in the study. 15613 households were selected for the study between 18th August and 20th September, 2020. A total of 29082 individuals provided whole blood samples and responded to a structured questionnaire. The authors used a chemiluminescent microparticle immunoassay to detect IgG antibodies against N protein of SARS-CoV-2. 3135 individuals tested seropositive. Among the seropositive individuals, 99 reported symptoms. The remaining 3029 (96.6%) were asymptomatic.

**XV. Iran[15] –** The authors investigated seroprevalence in the general population and in individuals with high risk of exposure across 18 cities in Iran starting from 17th April to 2nd June, 2020. Individuals were randomly selected from either the national healthcare registry or the employee lists of various agencies with high infection exposure risk. Venous blood was collected from the participants after their response to the questionnaire was recorded. An ELISA was used to detect SARS-CoV-2 specific IgG and IgM in the serum. Of a total of 3530 participants, who both got tested and responded to the questionnaire, 1164 tested seropositive. Of the latter, 416 declared not to have experienced symptoms.

**XVI. Ireland[16] –** The authors sent invitation letters in the week of 15th June 2020 to randomly selected individuals from a health service database from two counties in Ireland to participate in the seroprevalence study. A telephonic interview was taken from willing individuals based on a structured questionnaire. All recruited individuals were between 12 and 69 years of age. Whole blood samples were collected in the local clinic between 22nd June to 16th July 2020. A total of 1733 samples were collected for whom the personal data and the symptomatology were also available. The authors used an ELISA to detect anti-spike IgG, IgM, and IgA antibodies in the serum. 33 individuals were seropositive, among whom 24 participants reported experiencing symptoms.

**XVII. Italy[17] –** In a national serosurvey lasting from 25th May to 15th July, 2020, 64,660 individuals participated by providing blood samples and responding to a questionnaire. An ELISA was used to detect anti-SARS-CoV-2 IgG antibodies. 2.5% of the participants were seropositive. 27.3% of the seropositive individuals were asymptomatic.

**XVIII. Jordan[18] –** A nationwide seroprevalence study was carried out in three phases during August 2020, October 2020 and January 2021. Each phase lasted approximately one month. In each phase, approximately 5000 individuals across all governorates (administrative regions) of Jordan were randomly selected and recruited. We chose the data of the third phase because it had the highest number of seropositive samples. In this phase, 5044 individuals provided venous blood samples and responded to the questionnaire. An ELISA was used to detect SARS-CoV-2 specific IgG and IgM antibodies. 1723 individuals were seropositive, among whom 1071 declared not having experienced symptoms. Vaccination had not commenced in Jordan before the end of the study period.

**XIX. Lithuania[19] –** In a national seroprevalence survey spanning 10th August to 10th September 2020, six major municipalities were selected for randomly sampling the citizens from the state registers. A total of 3087 adult participants consented to provide capillary blood samples and responded to a questionnaire. The authors used a rapid immuno-chromatographic test to detect anti-SARS-CoV-2 IgG and IgM antibodies. Among the seropositive individuals, 69% reported not having experienced any symptoms.

**XX. Luxembourg[20] –** In a nationwide serosurvey, participants were randomly selected based on information about their age, gender and residency. Willing participants were enrolled from 15th April to 15th May, 2020. The cohort was stratified for age and sex to be representative of the general population of Luxembourg. Blood samples were self-collected by the participants using the kits they were provided. A total of 1807 samples were collected. Personal data and symptomatology were also obtained via a questionnaire. An ELISA was used to detect anti-SARS-CoV-2 IgG and IgA antibodies. 35 individuals were seropositive, of whom 10 individuals reported no symptoms.

**XXI. Mexico[21] –** A national serosurvey spanning August to November, 2020 randomly selected participants across nine administrative regions of the nation. A total of 9640 individuals consented to provide blood samples and responded to a questionnaire. The authors used an immunoassay to detect IgG antibodies against N proteins of SARS-CoV-2. Among the participants, 24.9% were seropositive, of whom 67.3% were asymptomatic.

**XXII. Netherlands[22] –** In a nationwide serosurvey, participants of a previous nationwide serosurvey which took place in 2017 to assess the seroprevalence against vaccine-preventable diseases were invited via post on 25th March, 2020. The willing individuals responded to an online questionnaire and consented to provide blood samples. The participants self-collected capillary blood by doing a finger prick using a kit which they received via postal service. The collection of the samples lasted from 31st March to 11th May, 2020. A total of 3147 samples were collected. The authors used a fluorescent bead based multiplex immunoassay to detect anti-spike IgG antibodies. 74 samples tested seropositive. 6.8% of the seropositive individuals reported not to have experienced any symptoms.

**XXIII. Oman[23] –** In a nationwide seroprevalence study, households were selected across all 11 governorates (administrative regions) of the nation by randomly choosing active telephone numbers to call and recruit individuals from households. Members within households were also selected randomly. Children under five years of age were excluded. The study was executed in four phases, lasting from July to November, 2020. Venous blood was collected from 17457 individuals. Anti-SARS-CoV-2 IgG antibodies were detected in the serum with a chemiluminescence assay. 3841 individuals were seropositive. Among them, 279 individuals declared to have experienced symptoms.

**XXIV. Pakistan[24] –** A seroprevalence study was conducted in the largest metropolitan city in Pakistan, Karachi, in three phases between April to August, 2020. The city was the epicenter of the pandemic in Pakistan. Different diasporas across Pakistan are well represented in the city. Households were randomly selected across different districts of the city. A total of 3005 participants provided blood samples and responded to a questionnaire. The authors tested the samples with an ELISA to detect anti-SARS-CoV-2 IgG and IgM antibodies. Among 364 seropositive participants, 333 (91.5%) individuals reported no symptoms.

**XXV. Peru[25] –** The study was carried out within the Lambayeque region, situated in northern Peru, between 24 June and 10 July, 2020. Participants aged nine and above were selected through multistage sampling. Initially, 38 districts were selected with probabilities proportional to their population size. This was followed by random sampling within each area in a district, and then household selection within the areas. Finally, individuals to be interviewed were chosen within the households. Individuals previously diagnosed with SARS-CoV-2 by RT-PCR or serological tests were excluded. Antibody positivity was determined using a lateral flow test. Of the 2,010 individuals surveyed, the seroprevalence was 29.5% (95% CI 27.6, 31.5), which was similar across the three provinces. 25.4% of these cases were asymptomatic.

**XXVI. Portugal[26] –** In a nationwide seroprevalence study spanning 21st May to 8th July, 2020, individuals were recruited across all regions of the nation including autonomous territories. Blood samples were collected from individuals older than 1 year who visited public hospitals or partnering private laboratories for causes unrelated to COVID-19. Individuals with medical conditions likely to interfere with the immune response were excluded. A total of 2301 individuals consented to provide blood samples and responded to a questionnaire. The samples were tested with two different ELISAs to detect anti-SARS-CoV-2 IgG and IgM antibodies respectively. 2.9% of the participants were seropositive, of whom 44% were asymptomatic.

**XXVII. Senegal[27] –** Representative clusters of households were selected across all 14 administrative regions in Senegal. A random selection was made to choose the households within the clusters and then individuals within the households. Between 25th October and 26th November, 2020, a total of 1422 individuals provided blood samples and completed a questionnaire. Anti-SARS-Cov-2 IgG/IgM antibodies against the S1/RBD viral proteins were detected in the serum. 398 individuals tested seropositive. Among them, 129 declared no symptoms.

**XXVIII. Spain[28] –** Between April 27 and June 22, 2020, a population-based nationwide serosurvey named ENE-COVID involved the participation of 61,092 community-dwelling individuals. These participants completed a questionnaire on symptoms and underwent an immunoassay to detect SARS-CoV-2 IgG antibodies. The selection of participants followed a two-stage sampling procedure, stratified by province and municipality size, from a pool of 88,653 individuals contacted. Among those who participated (comprising 68.9% of the contacted individuals), two serology tests, a point-of-care test, and a chemiluminescent microparticle immunoassay (CMIA) were used to identify the presence of SARS-CoV-2 antibodies. The prevalence of asymptomatic infections among the 2,669 individuals who tested seropositive was determined to be 28.7%.

**XXIX. Switzerland[29] –** A seroprevalence study was conducted in the Geneva canton (administrative region) of Switzerland. Participants were invited via email from a health survey database consisting of 20 to 74 year old individuals in the canton. These individuals were requested to enroll their household members who were more than five years old. The enrollment phase lasted from 6$^{th}$ April to 30$^{th}$ June, 2020. Blood samples from a total of 8,344 individuals were collected along with their responses to a questionnaire. An ELISA was used, in addition to another similar immuno fluorescence assay, to detect IgG antibodies against the S protein. 590 individuals were seropositive. Among the seropositive individuals, 77 (13%) participants declared not to have experienced any symptoms.

**XXX. UK[30] –** The Real-time Assessment of Community Transmission 2 (REACT-2) study was conducted in England from 20$^{th}$ June to 13$^{th}$ July, 2020 for evaluating the community prevalence of COVID-19. Individuals more than 18 years of age were randomly chosen from 315 lower-tier local authorities across England. Via post, willing participants received test kits for detecting IgG antibodies against SARS-CoV-2 in finger-pricked blood in a self-administered lateral flow immunoassay. A total of 1,05,651 individuals completed the questionnaire and the test. 5,544 participants were seropositive. Among the seropositive individuals, 3,759 individuals reported symptoms, so that 32.2% were asymptomatic.

**XXXI. USA[31] –** A cross sectional seroprevalence survey was conducted on adult participants (>18 y) recruited from all 50 states, the federal district, and from the overseas territories of USA. The enrollment lasted from March to August, 2020. The participants were tested longitudinally for the presence of anti-SARS-CoV-2 antibodies. A total of 4510 individuals underwent serological tests, starting from May 2020 and ending in January 2021. Dried blood sample collection kits were provided to the participants via the postal service for collection of capillary blood at home. These samples were analyzed using an ELISA to detect IgG, IgA, IgM antibodies against the N protein. Among the participants, 7.3% seroconverted. 28% of the seroconverted individuals reported not having experienced any symptoms. Although the study end date was slightly beyond our cutoff date for the systematic review, we included it in our meta-analysis because the authors mentioned in the report that their sample collection predated vaccine rollout.

**XXXII. Yemen[32] –** In November–December, 2020, a cross-sectional study encompassing 2001 participants from various age groups across four districts in Aden, southern Yemen was conducted. Employing a multi-stage sampling technique, data was collected through a structured questionnaire covering demographic and clinical variables. Blood samples were obtained from all participants. For testing, Healgen COVID-19 IgG/IgM Rapid Diagnostic Test (RDT) Cassettes were given to all participants. Subsequently, all positive RDT results and 14% of negative RDT outcomes underwent confirmation with the WANTAI SARS-CoV-2 Ab ELISA Kit. Of the 2001 participants, 549 were found to be RDT positive and confirmed by ELISA, indicating a prevalence of 27.4%. 157 seropositive individuals were found to be asymptomatic.

**XXXIII. Zambia[33] –** Seroprevalence was measured in six districts across Zambia starting on 4$^{th}$ July, 2020 and ending on 27$^{th}$ July, 2020. 16 areas were randomly chosen in each district, and 20 households were selected from these areas again at random. The participants provided nasopharyngeal swab samples for RT-PCR tests. Blood samples were also collected either by finger prick or heel prick (if less than 6 months old), or by venipuncture if preferred by the participant or when finger/heel puncture failed. An ELISA was used to detect IgG antibodies against the S protein. Participants also responded to a questionnaire on symptomatology. The total number of participants was 1886. Of them, 205 were seropositive. Of the latter, 23.8% reported symptoms.

### Text S3: Description of critical appraisal process

The guidelines by Joanna Briggs institute (JBI) for reviews on prevalence and incidence offer a critical appraisal checklist specifically designed for prevalence studies[34]. No such checklists exist for studies on the prevalence of asymptomatic infections. We therefore modified the JBI checklist and developed our own quality assessment tool and grading scale. Our critical appraisal checklist includes the following questions:

1.1 Was the sample frame appropriate to address the target population?

1.2 Were study participants sampled randomly?

1.3 Was the participation rate adequate (>70%)?

2.0 Were the study subjects and the setting described in detail?

3.1 Was there a clear definition for an asymptomatic case?

3.2 Was symptom assessment carried out in a standard, objective way?

Acceptable answers are ‘YES,’ ‘NO,’ and ‘UNCLEAR.’

Our quality assessment tool incorporates the domains of symptom assessment and random sampling method to ensure representativeness at a national level. Studies were classified as of low, moderate, or high quality following grading scales developed a priori: If the answer to question 1.2 is negative, the study is automatically of low quality. If one but not both of the questions assessing the validity of methods used to identify the clinical manifestation of infection (3.1, 3.2) is answered "no" or "unclear," then the study is of moderate quality. If both the questions (3.1 and 3.2) are answered "no" or "unclear," then study is of low quality.

**Table S1: Estimates of $\psi$ across independent studies from specific nations.** Studies not included in our meta-analysis are indicated using black squares. The other details are identical to those in Table 1.

| **Nation** | | **Study period** | **Cohort size** | **Seropo-sitive** | **Asympto-matic** | $\psi$ (95% CI) | **Q** | **G** |
|---|---|---|---|---|---|---|---|---|
| Austria[1] | | 21-Apr-20 to 27-Apr-20 | 1259 | 566 | 102 | 18.0 (15.1-21.4) | M | L |
| Austria[35] | ■ | 01-Jun-20 to 15-Jun-20 | 862 | 71 | 14 | 19.7 (12.1-30.4) | M | L |
| Chile[4] | | 25-Sept-20 to 25-Nov-20 | 2493 | 242 | 89 | 36.7 (31.0-43.0) | H | S |
| Chile[36] | ■ | 24-Apr-20 to 21-Jun-20 | 1368 | 93 | 18 | 18.9 (12.0-28.0) | M | L |
| China[5] | | 10-Apr-20 to 18-Apr-20 | 34857 | 427 | 326 | 76.3 (72.0-80.0) | H | N |
| China[37] | ■ | 14-Apr-20 to 15-Apr-20 | 9702 | 532 | 437 | 82.1 (78.6-85.1) | H | L |
| France[9] | | 26-Jun-20 to 24-Jul-20 | 2006 | 43 | 7 | 16.3 (8.1-30.0) | M | L |
| France[38] | ■ | 30-Mar-20 to 30-Apr-20 | 2004 | 306 | 56 | 18.3 (14.4-23.0) | L | L |
| Germany[11] | | 20-May-20 to 09-Jun-20 | 2203 | 249 | 61 | 24.5 (19.6-30.2) | M | L |
| Germany[39] | ■ | 13-May-20 to 22-May-20 | 620 | 52 | 13 | 25.0 (15.2-38.2) | L | L |
| India[14] | | 18-Aug-20 to 20-Sep-20 | 29082 | 3135 | 3029 | 96.6 (95.9-97.2) | M | N |
| India[40] | ■ | 01-Aug-20 to 31-Aug-20 | 4146 | 862 | 780 | 90.5 (88.3-92.3) | H | L |
| Spain[28] | | 27-Apr-20 to 22-Jun-20 | 61092 | 2669 | 766 | 28.7 (26.1-31.4) | M | N |
| Spain[41] | ■ | 27-Apr-20 to 11-May-20 | 61075 | 3054 | 999 | 32.7 (31.1-34.4) | H | N |

**Table S2: Critical appraisal of the studies included in the systematic review.** The column headings – 1.1, 1.2, etc. – refer to the questions in Text S3.

| Nation | 1.1 | 1.2 | 1.3 | 2.0 | 3.1 | 3.2 | Quality |
|---|---|---|---|---|---|---|---|
| Austria[1] | Yes | Yes | Yes | Yes | No | Yes | Moderate |
| Brazil[2] | Yes | Yes | Yes | Yes | Yes | Yes | High |
| Canada[3] | Yes | Yes | No | Yes | Yes | Yes | High |
| Chile[4] | Yes | Yes | Yes | Yes | Yes | Yes | High |
| China[5] | Yes | Yes | Yes | Yes | Yes | Yes | High |
| Colombia[6] | Yes | Yes | Unclear | Unclear | Yes | Yes | High |
| Denmark[7] | Yes | Yes | No | Yes | Yes | Yes | High |
| Ethiopia[8] | Yes | Yes | Unclear | Yes | Unclear | Yes | Moderate |
| France[9] | Yes | Yes | Unclear | Yes | Unclear | Yes | Moderate |
| Georgia[10] | Yes | Unclear | Unclear | Yes | Unclear | Yes | Low |
| Germany[11] | Yes | Yes | Unclear | Unclear | Unclear | Yes | Moderate |
| Ghana[12] | Yes | Unclear | Yes | Yes | Unclear | Yes | Low |
| Hungary[13] | Yes | Yes | Yes | Yes | Unclear | Yes | Moderate |
| India[14] | Yes | Yes | Yes | Yes | Unclear | Yes | Moderate |
| Iran[15] | Yes | Yes | Yes | Yes | Yes | Yes | High |
| Ireland[16] | Yes | Yes | No | Yes | Unclear | Yes | Moderate |
| Italy[17] | Yes | Yes | Unclear | Yes | Yes | Yes | High |
| Jordan[18] | Yes | Yes | Yes | Yes | No | No | Low |
| Lithuania[19] | Yes | Yes | No | Yes | Yes | Yes | High |
| Luxembourg[20] | Yes | Yes | Unclear | Yes | Yes | Yes | High |
| Mexico[21] | Yes | Yes | No | Yes | Yes | Yes | High |
| Netherlands[22] | Yes | Yes | Unclear | Yes | Unclear | Yes | Moderate |
| Oman[23] | Yes | Yes | Unclear | Yes | Unclear | Yes | Moderate |
| Pakistan[24] | Yes | Yes | Yes | Yes | Unclear | Yes | Moderate |
| Peru[25] | Yes | Yes | Unclear | Yes | Unclear | Yes | Moderate |
| Portugal[26] | Yes | Yes | Unclear | Yes | Yes | Yes | High |
| Senegal[27] | Yes | Yes | Yes | Yes | Unclear | Yes | Moderate |
| Spain[28] | Yes | Yes | No | Yes | No | Yes | Moderate |
| Switzerland[29] | Yes | Yes | No | Yes | Yes | Yes | High |
| UK[30] | Yes | Yes | Unclear | Yes | Yes | Yes | High |
| USA[31] | Yes | Unclear | Unclear | Yes | Unclear | Yes | Low |
| Yemen[32] | Yes | Yes | Unclear | Yes | Unclear | No | Low |
| Zambia[33] | Yes | Yes | Unclear | Yes | Unclear | Yes | Moderate |

**Table S3: Weights assigned to studies in the random-effects model.**

| Nation | Weight (%) | Nation | Weight (%) |
|---|---|---|---|
| Austria | 3.08 | Jordan | 3.1 |
| Brazil | 3.09 | Lithuania | 2.94 |
| Canada | 3.08 | Luxembourg | 2.9 |
| Chile | 3.08 | Mexico | 3.1 |
| China | 3.08 | Netherlands | 2.81 |
| Colombia | 3.1 | Oman | 3.1 |
| Denmark | 3.08 | Pakistan | 3.05 |
| Ethiopia | 3.03 | Peru | 3.09 |
| France | 2.86 | Portugal | 3.01 |
| Georgia | 2.54 | Senegal | 3.08 |
| Germany | 3.07 | Spain | 3.1 |
| Ghana | 3.07 | Switzerland | 3.08 |
| Hungary | 3.02 | UK | 3.1 |
| India | 3.09 | USA | 3.06 |
| Iran | 3.1 | Yemen | 3.09 |
| Ireland | 2.89 | Zambia | 3.06 |
| Italy | 3.1 | | |

**Table S4: Reported versus calculated confidence intervals on $\psi$.** The calculated confidence intervals were based on the Wilson score interval (see Methods).

| **Nation** | **$\psi$ (95% CI) reported** | **$\psi$ (95% CI) calculated** |
|---|---|---|
| Brazil[2] | 12.1 (10.1-14.5) | 12.1 (10.1-14.5) |
| France[9] | 16.3 (8.1-30.0) | 16.3 (6.8-30.7) |
| Spain[28] | 28.7 (26.1-31.4) | 28.7 (27.0-30.4) |
| Spain[41] | 32.7 (31.1-34.4) | 32.7 (30.2-35.4) |
| UK[30] | 32.2 (31.0-33.4) | 32.2 (31.0-33.4) |

**Table S5: Data of the predictors.** DMA is demographic median age; CVDR is the prevalence of cardiovascular disease per $10^5$ individuals; HDI is the human development index; and Age≥ 65 y is the percentage of the population older than 65 years.

| Nation | DMA[42] | CVDR[43] | HDI[44] | Age ≥ 65 y[42] |
|---|---|---|---|---|
| Austria | 42.6 | 11877.07 | 0.91 | 19.15 |
| Brazil | 32.4 | 5883.25 | 0.76 | 9.29 |
| Canada | 39.9 | 11771.44 | 0.93 | 18.02 |
| Chile | 34.5 | 6003.17 | 0.85 | 12.40 |
| China | 37.4 | 8460.08 | 0.76 | 12.60 |
| Colombia | 30.4 | 4773.92 | 0.76 | 8.47 |
| Denmark | 41.2 | 10669.84 | 0.95 | 20.05 |
| Ethiopia | 18.3 | 3288.15 | 0.5 | 3.13 |
| France | 41.4 | 10238.63 | 0.9 | 21.01 |
| Georgia | 36.3 | 12927.40 | 0.8 | 14.50 |
| Germany | 44.9 | 11977.15 | 0.94 | 21.96 |
| Ghana | 20.2 | 4129.86 | 0.63 | 3.41 |
| Hungary | 42.5 | 14638.53 | 0.85 | 20.10 |
| India | 27.3 | 5114.69 | 0.64 | 6.67 |
| Iran | 31.4 | 8026.21 | 0.78 | 7.11 |
| Ireland | 37.3 | 8072.07 | 0.94 | 14.54 |
| Italy | 46.4 | 15937.50 | 0.89 | 23.37 |
| Jordan | 23.1 | 4097.09 | 0.72 | 3.61 |
| Lithuania | 43.5 | 14000.45 | 0.88 | 20.42 |
| Luxembourg | 38.6 | 9110.47 | 0.92 | 14.56 |
| Mexico | 28.7 | 4706.57 | 0.76 | 8.02 |
| Netherlands | 41.7 | 10305.62 | 0.94 | 19.65 |
| Oman | 28.8 | 3732.08 | 0.83 | 2.76 |
| Pakistan | 20.0 | 3850.80 | 0.54 | 4.17 |
| Peru | 28.0 | 4129.31 | 0.76 | 8.25 |
| Portugal | 44.7 | 10218.49 | 0.86 | 22.30 |
| Senegal | 17.6 | 3968.73 | 0.51 | 3.17 |
| Spain | 43.5 | 9820.96 | 0.90 | 19.67 |
| Switzerland | 41.7 | 9373.02 | 0.96 | 18.73 |
| UK | 39.5 | 9668.34 | 0.92 | 18.72 |
| USA | 37.5 | 12095.06 | 0.92 | 16.22 |
| Yemen | 18.5 | 3969.17 | 0.46 | 2.72 |
| Zambia | 16.8 | 3359.27 | 0.57 | 1.73 |

**Table S6: Model characteristics and parameter estimates**

**S6A: Model with HDI as predictor**

*All nations included*

| | Estimate (95% CI) | p-value |
|---|---|---|
| Intercept | $4.1\ (1.9, 6.4)$ | $4 \times 10^{-4}$ |
| HDI | $-5.5\ (-8.3, -2.6)$ | $2 \times 10^{-4}$ |
| $R^2$ | $30.0\%$ | |

*Excluding Oman, Senegal, and Yemen*

| | | |
|---|---|---|
| Intercept | $6.9\ (4.9, 8.8)$ | $4 \times 10^{-12}$ |
| HDI | $-8.6\ (-11.1, -6.4)$ | $4 \times 10^{-13}$ |
| $R^2$ | $65.5\ \%$ | |

**S6B: Model with DMA as predictor**

*All nations included*

| | Estimate (95% CI) | p-value |
|---|---|---|
| Intercept | $2.6\ (1.0, 4.2)$ | $1.4 \times 10^{-3}$ |
| DMA | $-0.1\ (-0.13, -0.04)$ | $4 \times 10^{-4}$ |
| $R^2$ | $27.3\ \%$ | |

*Excluding Senegal and Yemen*

| | | |
|---|---|---|
| Intercept | $3.8\ (2.1, 5.4)$ | $1 \times 10^{-5}$ |
| DMA | $-0.1\ (-0.2, -0.1)$ | $2 \times 10^{-6}$ |
| $R^2$ | $42.4\ \%$ | |

**S6C: Model with CVDR as predictor**

*All nations included*

| | Estimate (95% CI) | p-value |
|---|---|---|
| Intercept | $1.2\ (0.2, 2.3)$ | $2 \times 10^{-2}$ |
| CVDR | $-2 \times 10^{-4}$ $(-3 \times 10^{-4}, -1 \times 10^{-4})$ | $4 \times 10^{-3}$ |
| $R^2$ | $18.9\ \%$ | |

*Excluding Lithuania*

| | | |
|---|---|---|
| Intercept | $1.4\ (0.4, 2.5)$ | $9 \times 10^{-3}$ |
| CVDR | $-2 \times 10^{-4}$ $(-3 \times 10^{-4}, -1 \times 10^{-4})$ | $9 \times 10^{-4}$ |
| $R^2$ | $24.8\ \%$ | |

**Table S7: Sensitivity and specificity of the assays used.**

| Nation | Test manufacturer | Test type | Sensitivity | Specificity |
|---|---|---|---|---|
| Austria[1] | EUROIMMUN | ELISA | N/A* | N/A |
| Brazil[2] | Guangzhou Wondfo Biotech Co. Ltd | LFIA | 84.80% | 99.95% |
| Canada[3] | National Microbiology Laboratory of Canada | CLIA | 98.00% | 99.00% |
| Chile[4] | Roche Diagnostics,Zhuhai Livzon Diagnostics Inc | ELISA, point of care | 90.6% to 99% | 99.8% to 99.2% |
| China[5] | Guangzhou Wondfo Biotech Co. Ltd,Innovita Biological Technology Co. Ltd,Bioscience Diagnostics,NA | Multiple Types | 76% | 100%, 98-100%, 100% |
| Colombia[6] | Eurofins Ingenasa | ELISA | 92.5 | N/A |
| Denmark[7] | Beijing Wantai Biological | ELISA | 96.70% | 99.50% |
| Ethiopia[8] | Abbott Laboratories | CLIA | 54.50% | 100.00% |
| France[9] | Bio-rad, Biosynex, EUROIMMUN | ELISA | N/A | N/A |
| Georgia[10] | Zhejiang Orient Gene Biotech | LFIA | 93.10% | 99.20% |
| Germany[11] | EUROIMMUN | ELISA | 88.30% | 99.20% |
| Ghana[12] | Wuhan UNscience Biotechnology Co. Ltd | LFIA | 66.00% | 94.00% |
| Hungary[13] | Abbott Laboratories | CLIA | 87.20% | N/A |
| India[14] | Abbott Laboratories | CLIA | 100.00% | 99.60% |
| Iran[15] | Pishtaz Diagnostics Iran | ELISA | 66.90% | 98.20% |
| Ireland[16] | Abbott Laboratories,Beijing Wantai Biological | Multiple Types | 93.90% | 100% |
| Italy[17] | Abbott Laboratories | CLIA | >= 90 % | N/A |
| Jordan[18] | Beijing Wantai Biological | ELISA | N/A | N/A |
| Lithuania[19] | AMEDA Labordiagnostik GmbH | LFIA | 92.00% | 99.40% |
| Luxembourg[20] | EUROIMMUN | ELISA | 85.70% | 97.80% |
| Mexico[21] | Roche Diagnostics | CLIA | 92.02% | 99.52% |
| Netherlands[22] | N/A | ELISA | 84.40% | 99.00% |
| Oman[23] | DiaSorin | CLIA | N/A | N/A |
| Pakistan[24] | Roche Diagnostics | CLIA | 100.00% | 99.80% |
| Peru[25] | Core Technology Co. Ltd | LFIA | 66% | 97% |
| Portugal[26] | EUROIMMUN, Beijing Wantai Biological | ELISA | 86.10% | 99.40% |
| Senegal[27] | Generic Diagnostics Ltd, Beijing Wantai Biological | ELISA | 84% to 100% | 67% to 100% |
| Spain[28] | Abbott Laboratories | CMIA | 90.6% | 99.3% |
| Switzerland[29] | Biotech | ELISA | 93.00% | 100.00% |
| UK[30] | Fortress Diagnostics | LFIA | 84.40% | 98.60% |
| USA[31] | Bio-rad | ELISA | 91.70% | 98.80% |
| Yemen[32] | Healgen,Beijing Wantai Biological | Multiple Types | N/A | N/A |
| Zambia[33] | EUROIMMUN | ELISA | 90% | N/A |

*N/A: Not available

**Table S8: Reported seropositivity in children.**

| Nation | Age range | Seropositivity (%) |
|---|---|---|
| Brazil[2] | 0-9y | 7.3 |
| China[5] | 1-9y | 0.7 |
| Jordan[18] | 0-9y | 0.4 |
| Portugal[26] | 1-9y | 2.9 |
| Zambia[33] | 0-9y | 4.0 |

**Table S9: Model characteristics and parameter estimates with age≥65 y as predictor.** Meta-regression of $\psi$ on percentage of the population with age ≥ 65 years. Yemen was an influential study following the criteria described in Figs. S3 and S8.

*All nations included*

| | **Estimate (95% CI)** | **p-value** |
|---|---|---|
| Intercept | $1.3\ (0.2,\ 2.1)$ | $3 \times 10^{-3}$ |
| Age ≥ 65 | $-0.\ 11(-0.17,\ -0.06)$ | $10^{-4}$ |
| $R^2$ | $31\ \%$ | |

*Excluding Yemen*

| | | |
|---|---|---|
| Intercept | $1.5\ (0.6,\ 2.3)$ | $7 \times 10^{-4}$ |
| Age ≥ 65 | $-0.\ 13(-0.18,\ -0.07)$ | $2 \times 10^{-5}$ |
| $R^2$ | $37\ \%$ | |

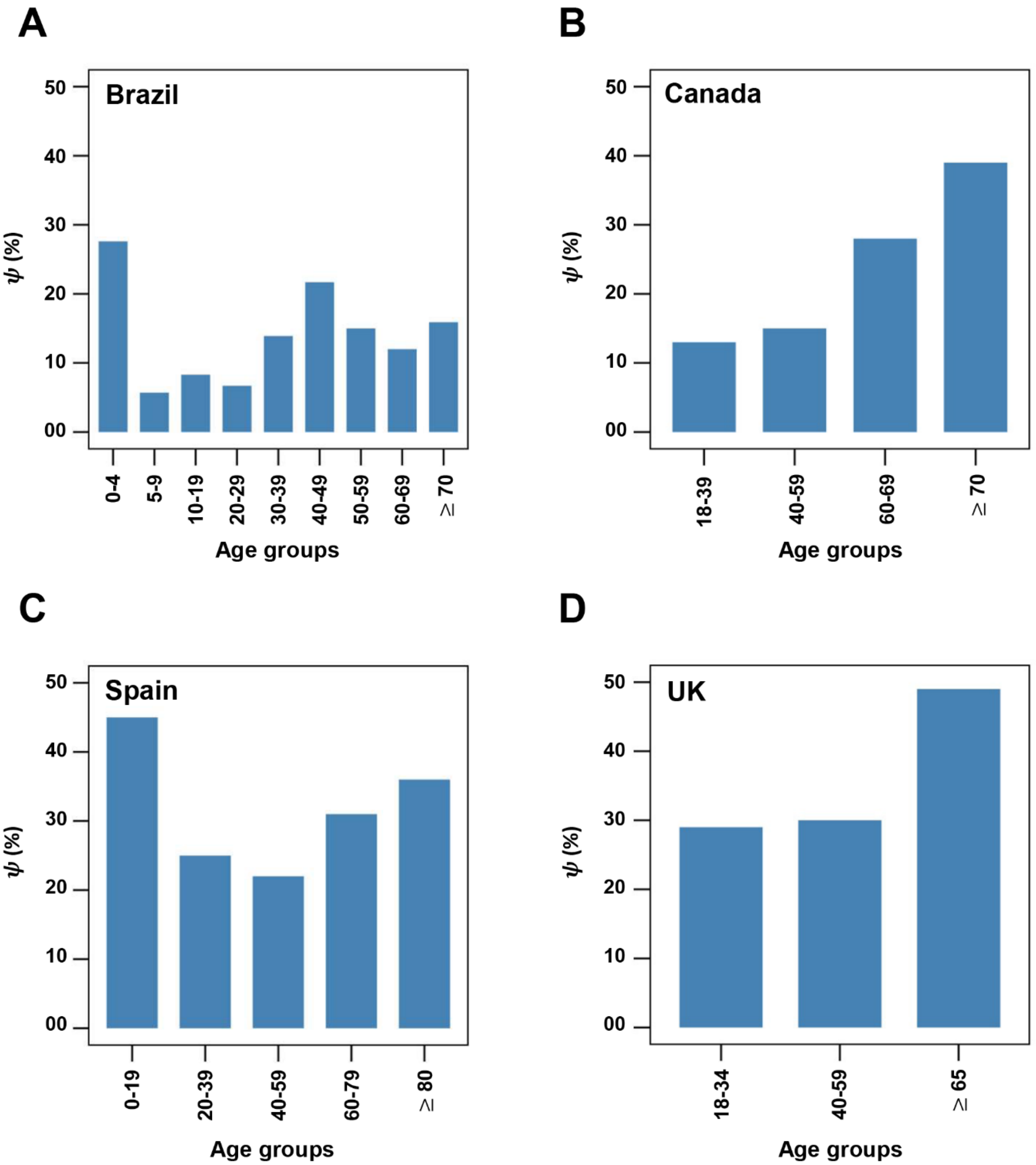


**Figure S1: Age stratified $\psi$ in studies of Brazil, Canada, Spain, and UK.** $\psi$ in different age groups was available for the studies from Brazil[2], Canada[3], Spain[28], and UK[30], which are reproduced above as bar charts. Age groups mentioned are the same as in the respective studies. Age is in years.

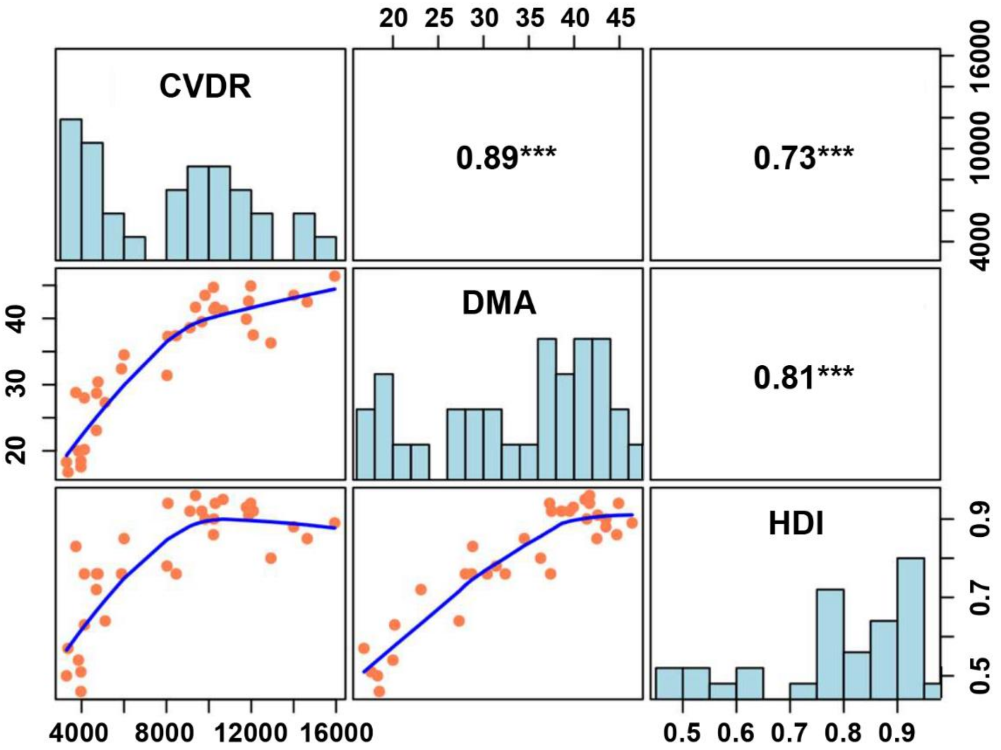


**Figure S2: Correlation between predictors.** Diagonal boxes show the distribution of each predictor across the 33 nations included. Sub-diagonal boxes show bivariate scatter plots with smooth fits (blue lines). Super-diagonal boxes show Spearman correlation coefficients between the predictors. ***: p<0.001.

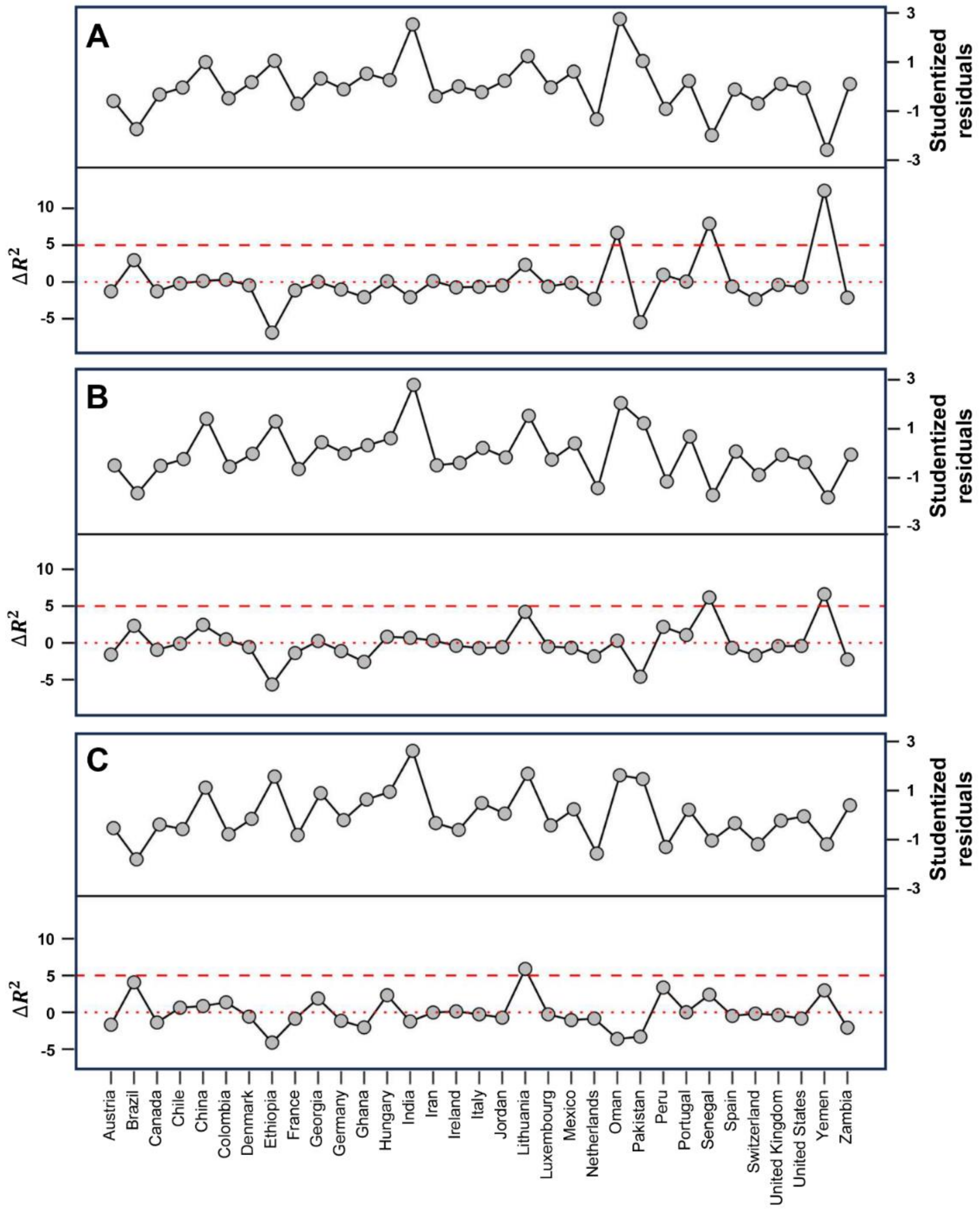


**Figure S3: Influential case diagnostics.** Plots of studentized residuals and $\Delta R^2$ when each study was removed in turn for the model with HDI (**A**), DMA (**B**), or CVDR (**C**) as predictor. $\Delta R_i^2 = R_i^2 - R_0^2$, where $R_i^2$ is the $R^2$ (indicated by the dashed red lines) when a specific study, denoted as '$i$,' is removed, and $R_0^2$ (indicated by the dotted red lines) is the $R^2$ when all the 33 studies were included. Outliers are identified by an absolute studentized residual exceeding 3, while studies with $\Delta R_i^2 \geq 5$ are considered influential.

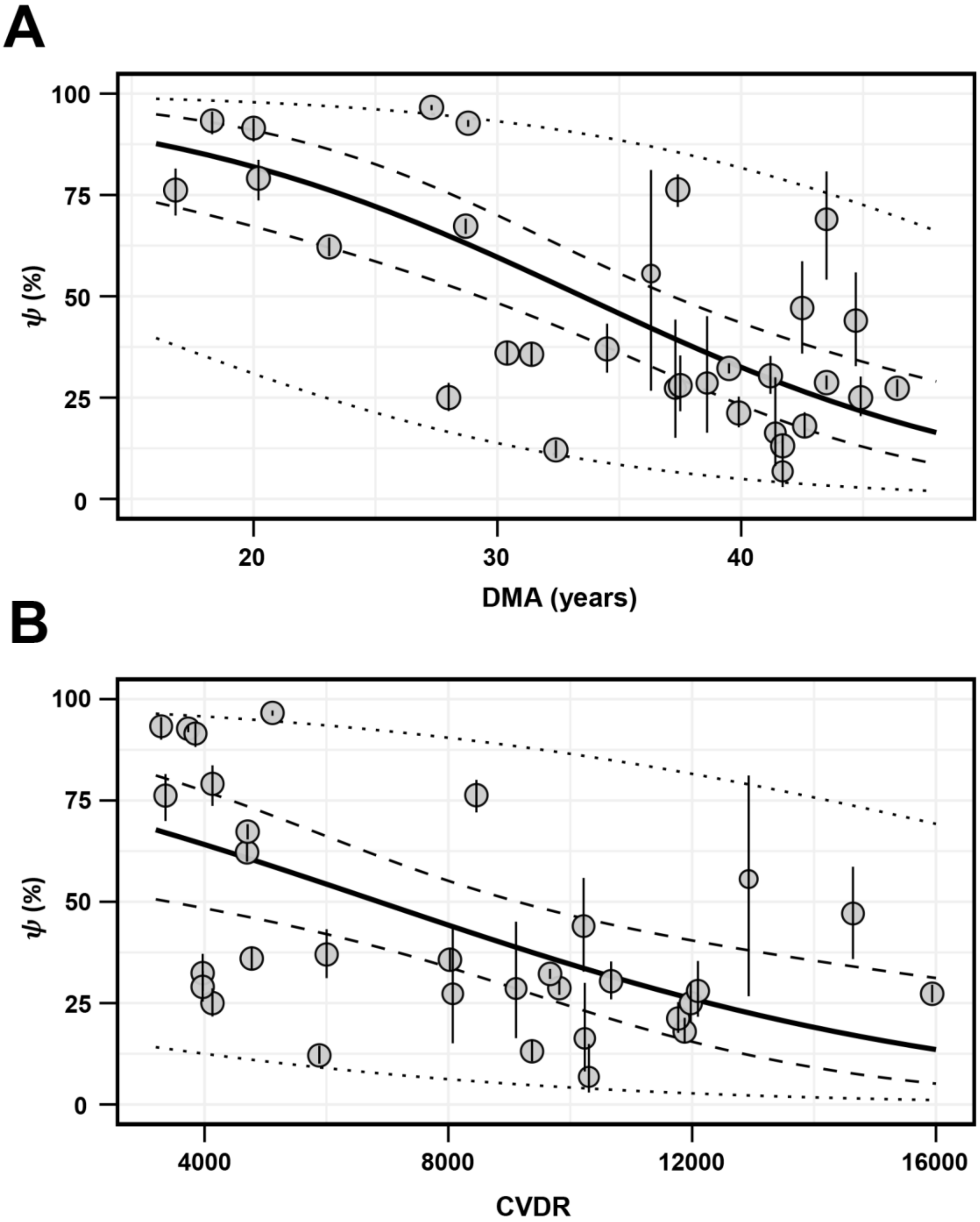


**Figure S4: Meta-regression with other predictors.** Best-fit meta-regression lines (solid line) showing the dependence of $\psi$ on DMA (**A**) and CVDR (**B**), overlayed on the estimates of $\psi$ (grey bubbles). The symbol sizes are proportional to the weights of the studies in the meta-regression (see Methods). Dashed lines are the 95% confidence intervals, and the dotted lines are the 95% prediction intervals (see Methods). Error bars are the same as in Fig. 2. Regression coefficients and statistics are in Table S6B and S6C. Influential studies as identified in Fig. S3 were excluded.

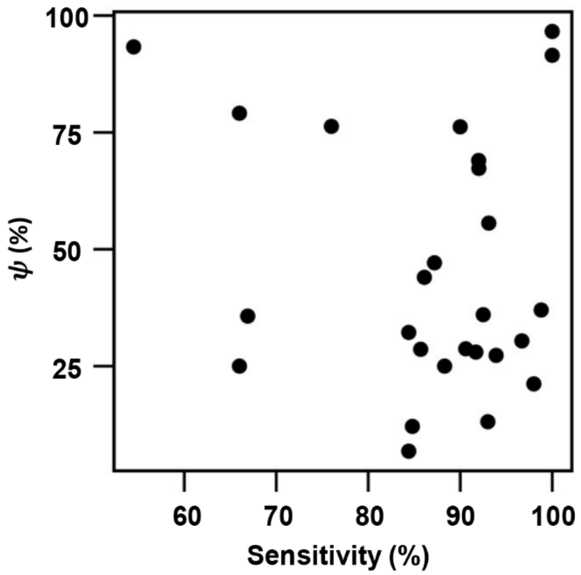


**Figure S5: Robustness to assay sensitivity.** Scatter plot between $\psi$ and the sensitivity of the serological assay used in the 33 studies included. Spearman correlation coefficient was 0.05 (p= 0.81).

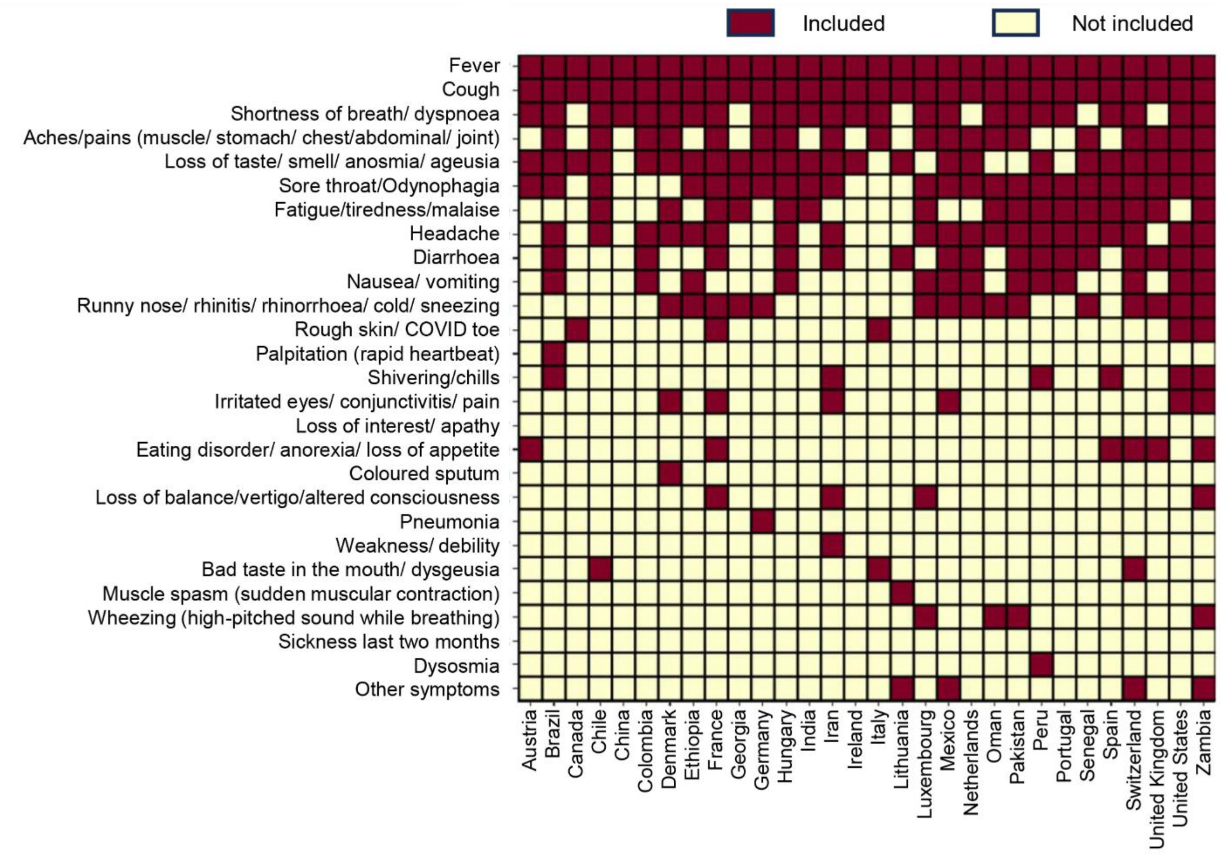


**Figure S6: Symptom sets.** Symptom sets used by the different studies in our meta-analysis. Studies from Jordan[18], Ghana[12], and Yemen[32] did not provide these details and are not included here.

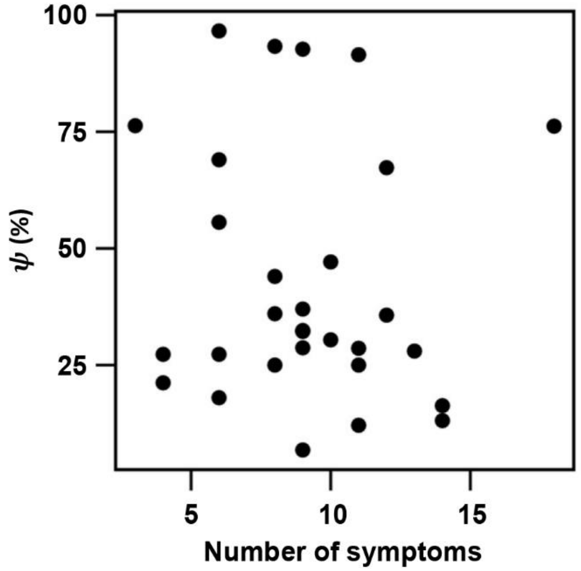


**Figure S7: Robustness to symptom sets.** Scatter plot between $\psi$ and the number of symptoms used in each of the studies included. Spearman correlation coefficient was −0.16 (p=0.41).

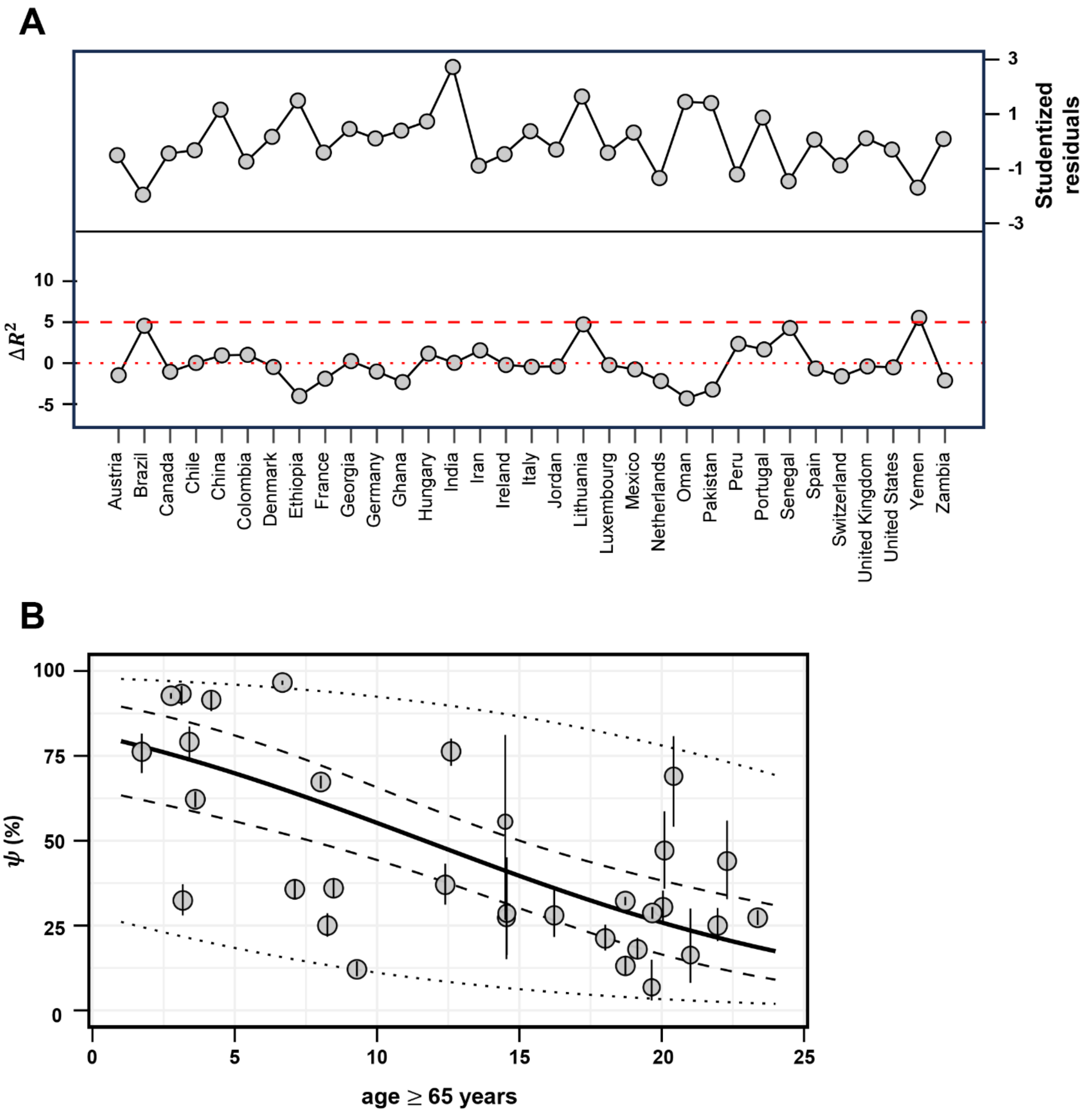


**Figure S8:** **Meta regression of $\psi$ on age $\geq$ 65 y. (A)** Influential case diagnosis as performed in Fig. S3. Yemen emerged as influential. **(B)** Meta-regression after excluding Yemen. Best-fit (solid line) showing the dependence of $\psi$ on age $\geq$ 65 years overlayed on the estimates of $\psi$ from 32 nations (grey bubbles). The symbol sizes are proportional to the weights of the studies in the meta-regression (see Methods). Dashed lines are the 95% confidence intervals and the dotted lines are the 95% prediction intervals. Error bars are the same as in Fig. 2. Regression coefficients and statistics are in Table S9.
